# Design and Implementation of PharMe: A Mobile Application to Return Pharmacogenomic Test Results to Patients

**DOI:** 10.1101/2025.06.10.25328968

**Authors:** Tamara Slosarek, Jannis Baum, Benjamin Frost, Nikola Genchev, Leon Hermann, Samuel Kunst, Charlie Meister, Matti Schmidt, Sebastian Wagner, Susan M. Mashni, Christoph Lippert, Erwin P. Böttinger, Aniwaa Owusu Obeng

## Abstract

**Objective:** Pharmacogenomics can optimize therapeutic outcomes while minimizing adverse events. However, barriers like the lack of trained healthcare professionals impede its widespread adoption. To address this challenge, we developed the PharMe app to return pharmacogenomic results to patients, facilitated by Anni, a custom annotation interface.

**Materials and Methods:** All components were developed user-centered; patient content was created by experts with Anni, based on CPIC and FDA guidelines. PharMe’s usability was tested in a remote pilot, asking users to perform tasks and give SUS and star ratings. A power user delivered in-depth, qualitative insights.

**Results:** PharMe is available for iOS and Android in English and includes information for 17 genes and 159 medications. Anni allows the consistent, multi-lingual annotation of guidelines with patient-friendly formulations. PharMe securely loads laboratory results, displays personalized annotated guidelines, and includes phenoconversion. Pilot testers finished most tasks successfully; on average, PharMe received 4.1 stars and a SUS score of 82.86. Feedback collected in all test sessions was incorporated into the current PharMe version.

**Discussion:** While the remote pilot testing has limitations like a small sample size and (digitally) literate participants, it showed that PharMe is a valuable, understandable, and easy-to-use system. PharMe is assessed in a clinical validation study in a more diverse population. In the future, PharMe and Anni could support further guidelines and data formats.

**Conclusion:** We developed a novel and user-friendly system to return pharmacogenomic results to patients and scale the clinical adoption of pharmacogenomics, supporting trained clinical experts. PharMe and Anni are designed to use transparent and standardized content based on published guidelines, be agnostic to pharmacogenomic data sources, and be flexibly deployed globally in different settings and institutions.

## Introduction

Adverse drug reactions are among the top ten leading causes of death and disease in developed countries.[1,2] For about 50 percent of the population, some commonly prescribed medications are ineffective.[3] Pharmacogenomics (PGx) has the potential to improve medication efficacy and safety significantly and can optimize therapeutic outcomes while minimizing adverse events.[4–6] However, many barriers impede its widespread adoption, such as the lack of trained healthcare professionals to adopt PGx into routine practice.[5,6] For example, at the Mount Sinai Health System in New York, consisting of eight hospital campuses with more than 9,000 physicians,[7] as of January 2025, only a dozen clinical pharmacists are trained to counsel patients following PGx testing. Additionally, Haga et al. [8] note that while patients generally view PGx testing positively and consider it valuable, the results must be effectively communicated by providers and made shareable across healthcare institutions.

To address this challenge, we developed PharMe, a mobile application designed to deliver PGx results directly to patients, supported by Anni, a custom-built, novel annotation interface. This approach aims to expand the practical use of PGx testing by reducing the need for time-intensive, in-person counseling with trained PGx experts.

Mobile applications have already been shown to improve patient outcomes in nutrition, cardiology, mental health, chronic disease management, and physical therapy.[9–12] In addition, digital tools have proven effective in communicating genetic results to patients.[13,14] While several services exist for returning PGx results in pharmacogenomics, most are designed for healthcare professionals rather than lay users,[15] such as PharmGKB’s DNA-Driven Rx tool [16]. Lipschultz et al. [17] implemented an administrative dashboard for generating patient summaries linked to a mobile-optimized, web-based patient portal. This portal displayed the summaries along with a traffic light system (red, yellow, green) and was integrated into an existing clinical decision support system. In evaluations of the portal, Truong et al. found that the portal access improved patients’ knowledge, understanding, and perceptions of PGx.[18] Patients also expressed a strong preference for a well-designed, secure portal that could be accessed via mobile devices and easily shared with their healthcare providers.[19] Blagec et al. [20] proposed the Medication Safety Code (MSC) system – a QR code representing patients’ PGx profile that can, e.g., be printed on a personalized card the size of a credit card and be kept in patients’ wallets. Healthcare providers can then scan a patient’s card and find information personalized for the patient in a web-based portal. As part of the U-PGx project, the PREPARE study utilized cards based on the MSC system to support clinical implementation of PGx across Europe, addressing challenges related to interoperability and data exchange between institutions.[21,22] Also, several commercial direct-to-consumer (DTC) testing companies offer PGx testing, although coverage of genetic variants and the interpretation guidelines used can vary significantly.[23] For example, 23andMe [24] began offering FDA-approved PGx reports in 2018 [25] without requiring physician consultation. These reports cover genes such as CYP2C19, DPYD, and SLCO1B1, as well as associated medications like clopidogrel, citalopram, and simvastatin. In contrast, companies like Pharmazam [26] Veritas [27] and Color [28] offer direct-to-consumer PGx testing through a hybrid model that involves physician ordering.

With PharMe, we aimed to develop a mobile platform for returning PGx test results to patients in a transparent, user-friendly format. The app is designed to present information that is easy to understand, grounded in published clinical guidelines, and compatible with a variety of PGx data sources – making it adaptable across different healthcare settings and institutions. Anni, our annotation interface, was developed to support creating patient-facing content by leveraging published guidelines, promoting consistency through reusable phrasing, and enabling multilingual support. This paper presents the design and implementation of the PharMe app and the Anni interface, along with initial findings from a usability evaluation.

## Materials and Methods

### Creation of User-Facing PGx Guideline Annotations

To improve the accessibility of expert-facing PGx guideline content, we developed annotations with patient-friendly language to enhance understanding among lay users. In the initial phase, A.O.O. – a PGx expert with experience in PGx counseling – collaborated with two final-year PharmD students to curate a spreadsheet. This spreadsheet included roughly 100 drug-gene pairs listed in rows, with associated phenotypes drawn from various guidelines arranged in columns. The curation and review process took a considerable amount of time, most of it searching for data from CPIC and other guideline sources. Moreover, the data was highly redundant and included many typographical errors and inconsistencies in formulations due to human error. Maintaining and updating the original dataset was challenging due to its complex structure and large number of columns, which made editing cumbersome. Additionally, the format only allowed data entry for single drug-gene pairs, making it unsuitable for representing guidelines involving multiple gene-phenotype combinations. The two-dimensional table layout also lacked the flexibility to accommodate multilingual annotations, which required manual translation. To address these limitations, we developed Anni through close collaboration between software engineers and PGx experts. We reconstructed and updated guideline annotations in Anni using the information from the original spreadsheet and implemented a formal review and publication workflow for the new annotations. Anni incorporates guideline information from the Clinical Pharmacogenomics Implementation Consortium (CPIC) and the U.S. Food and Drug Administration (FDA). General drug information was obtained from publicly available resources such as drug labels and DrugBank [29].

### PharMe Design and Prototype Development

An initial PharMe app prototype was created as part of a student project at Hasso Plattner Institute in Potsdam, Germany, from August 2021 to July 2022, in collaboration with the Mount Sinai Health System. We initially created a PretoVid^‡^ [30], which summarizes the rationale behind PharMe to assess preliminary user and expert feedback and attitudes towards the tool. Based on the findings, we designed app-wireframes and evaluated those again with users and experts (all interview guides are available in Supplementary Material S1). The initial app prototype was then developed based on this feedback and comprised most of the functions and design elements available in the current version. Preliminary validation of PharMe was conducted during a World Café [31] format workshop in Oldenburg, Germany, to evaluate PharMe’s usability and perceived usefulness (see Supplementary Material S2 for the discussion guide). This workshop was conducted as part of the Health-X dataLOFT project [32]. To further assess the app’s general usability and functionality, a remote pilot test was carried out with diverse users from Germany and the US, including colleagues, friends, and acquaintances. Participants received instructions via email, including a survey link to Google Forms (see Supplementary Material S3), credentials to access test data, and installation instructions for PharMe via TestFlight (iOS) or a Firebase test link (Android). The survey was based on the World Café guide and instructed testers to execute different tasks using PharMe and to provide feedback for every screen. After testing, participants were asked to rate the app with a short explanation of their rating and complete the System Usability Scale (SUS) [33]. Five-point Likert-type scale answers are converted to SUS scores ranging from 0 to 100; SUS scores were interpreted using the Sauro-Lewis curved grading scale (CGS) [34]. After the remote pilot testing exercise, we collected app feedback from more in-depth power user testing. We shared the app with a New York-based, 70- to-80-year-old Mount Sinai patient who had conducted PGx testing before and has a background in testing digital products. The tester followed the remote pilot testing protocol and explored the app independently beyond the protocol. We collected her feedback via email and a video call. The current version of the app being described here has suggestions and input from the user testing sessions incorporated.

## Results

### Included Guideline Data

To date, PharMe includes information for 17 genes and 159 medications (see Supplementary Material S5). This includes drug-gene pairs from CPIC guidelines [35] with level A or B (“prescribing action recommended” [36]) and FDA Table of Pharmacogenetic Associations [37] for medications that are not already included in CPIC guidelines with “data support for therapeutic management recommendations” or “potential impact on safety or response”. Notably, we only include guidelines for genes supported by the CPIC application programming interface (API) [38], namely RYR1, CYP2D6, TPMT, CACNA1S, SLCO1B1, G6PD, CYP3A5, HLA-B, CFTR, ABCG2, CYP2C19, DPYD, MT-RNR1, UGT1A1, CYP2B6, HLA-A, CYP2C9, and NUDT15. Moreover, for the initial app version being validated in the ongoing study, we excluded genes not covered in the AccessDx PGx Profile [39], i.e., RYR1, CACNA1S, CFTR, and MT-RNR1. Additionally, we have included CPIC level C (“published studies at varying levels of evidence […] but no prescribing actions are recommended” [36]) medications; seven CYP2D6 inhibitors without guidelines, namely bupropion, quinidine, terbinafine, abiraterone, cinacalcet, lorcaserin, and mirabegron; and three CYP2D6 inhibitors with guidelines, i.e., fluoxetine, paroxetine, and duloxetine, to enable phenoconversion. The information in the app reflects CPIC and FDA resources available as of May 16, 2025.

### Software Components

Figure 1 depicts the software architecture components and the data flow between them: the Annotation Interface (Anni), PharMe, a patient health data source, and Scio, an additional module to assess user comprehension. All PharMe-specific components are included in the open-source PharMe mono-repository on GitHub^§^.

**Figure 1.**
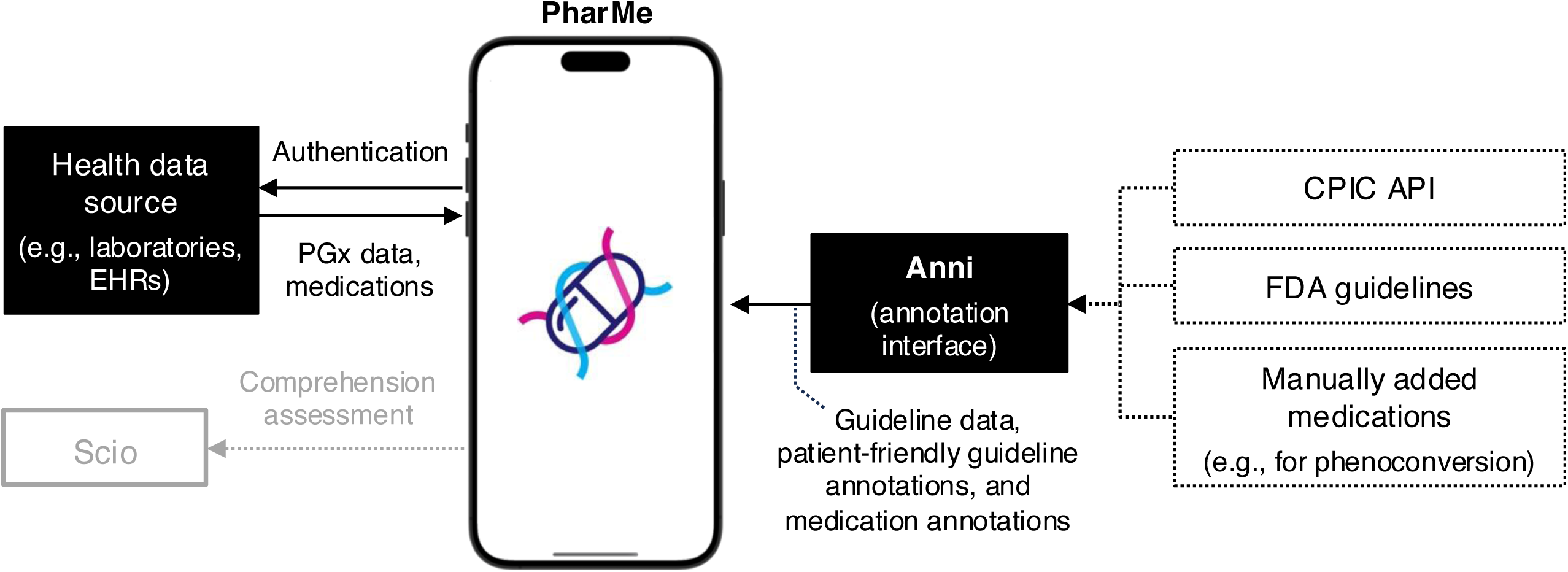
Architecture and data flow of PharMe, Anni, and a patient health data source.

#### Annotation Interface (Anni)

PGx experts can use Anni to view and annotate medications based on published PGx guidelines, utilizing pre-defined text bricks that can be made available within the app (Figure 2). Anni is a web-based application built with Next.js [40] and a MongoDB [41] document storage, providing a REST API to query the current annotation version and data.

**Figure 2.**
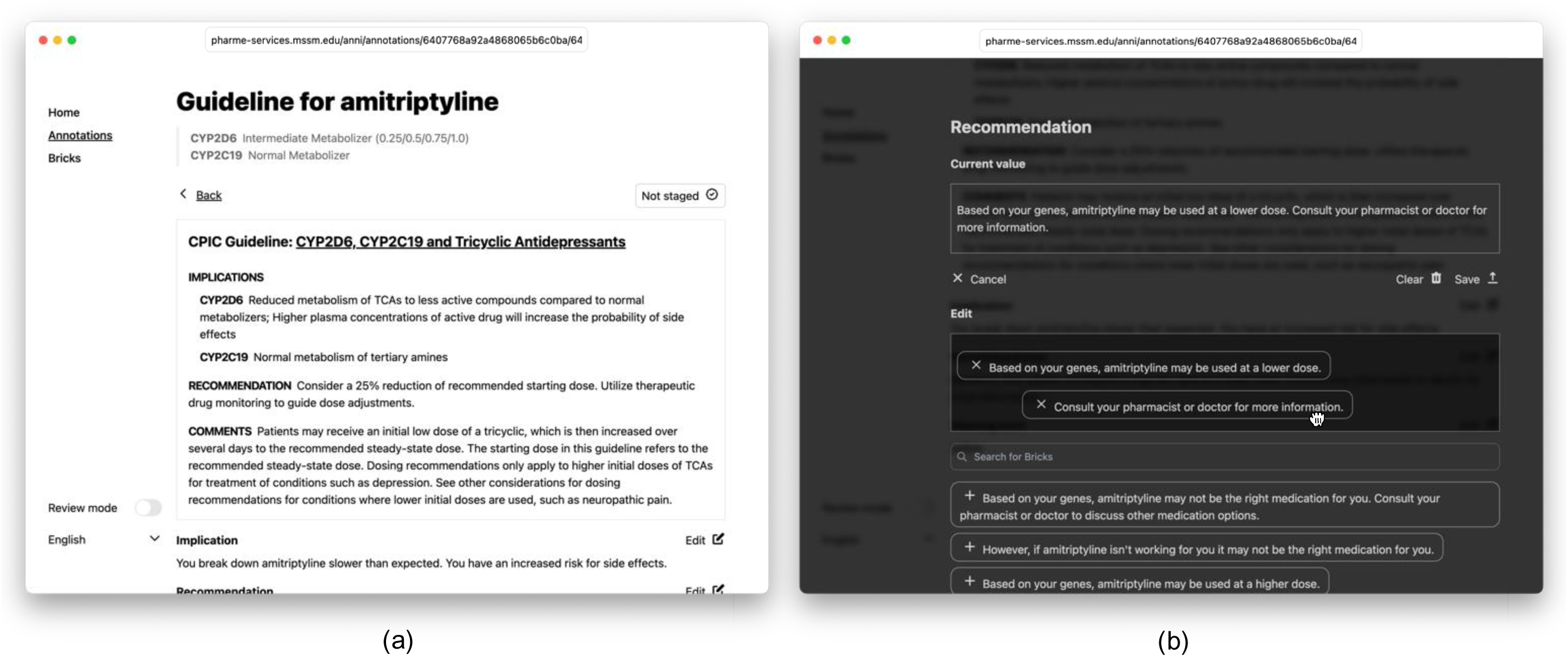
Annotation creation in Anni; (a) guideline data and annotations for one phenotype; (b) editing annotations with text bricks.

Medications and corresponding guidelines were uploaded using the CPIC API and a custom repository** for additional guidelines, such as the FDA table. For each phenotype of a gene-drug combination, annotations for the clinical implication, PGx-guided recommendation, and warning level (green, yellow, red, none) can be added while viewing the corresponding guideline (see Figure 2a, Supplementary Figure S1d). Anni also supports medication-level annotations with patient-friendly descriptions of drug class, intended use (indication), and common brand names (see Supplementary Figure S1b, Supplementary Figure S1c). All annotations consist of pre-defined, multi-lingual text bricks that can be selected using drag-and-drop (see Figure 2b, Supplementary Figure S1e, Supplementary Figure S1f). The text brick approach is designed to improve consistency and annotation speed, enable cross-checking across medications and guidelines, and significantly reduce the effort required to translate annotations into other languages. After annotations are added and edited, a review step is introduced: the author or another expert can activate the “review mode” in Anni, which allows annotations to be staged for publishing. In this mode, editing is disabled to avoid accidental changes. After review, staged annotations can be published and made available for PharMe (see Supplementary Figure S1a). This three-part process aims to ensure the quality and integrity of the annotations and to offer a development environment where one can edit the annotations without directly changing what is shown to end-users in PharMe. Moreover, Anni can be launched with a backup service, and it includes update scripts that compare the latest guideline data with the current data in Anni.

#### PharMe

PharMe is a mobile application developed with the multi-platform framework Flutter [42] that displays PGx test results to patients, using patient-friendly content from clinical PGx guidelines curated in Anni (Figure 3, Supplementary Figure S2a). After successful authentication, the app receives patients’ genetic and medication data from a patient health data source (see Supplementary Figure S2b). PharMe is designed to be agnostic to the data source, and the data format and authentication methods can be implemented individually. In addition, PharMe downloads all annotation data from Anni and selects the guidelines matching the users’ genetic data using CPIC lookup keys (see Figure 4). The matching is done solely on the phone, and all data is stored in encrypted form to ensure data privacy and security.

**Figure 3.**
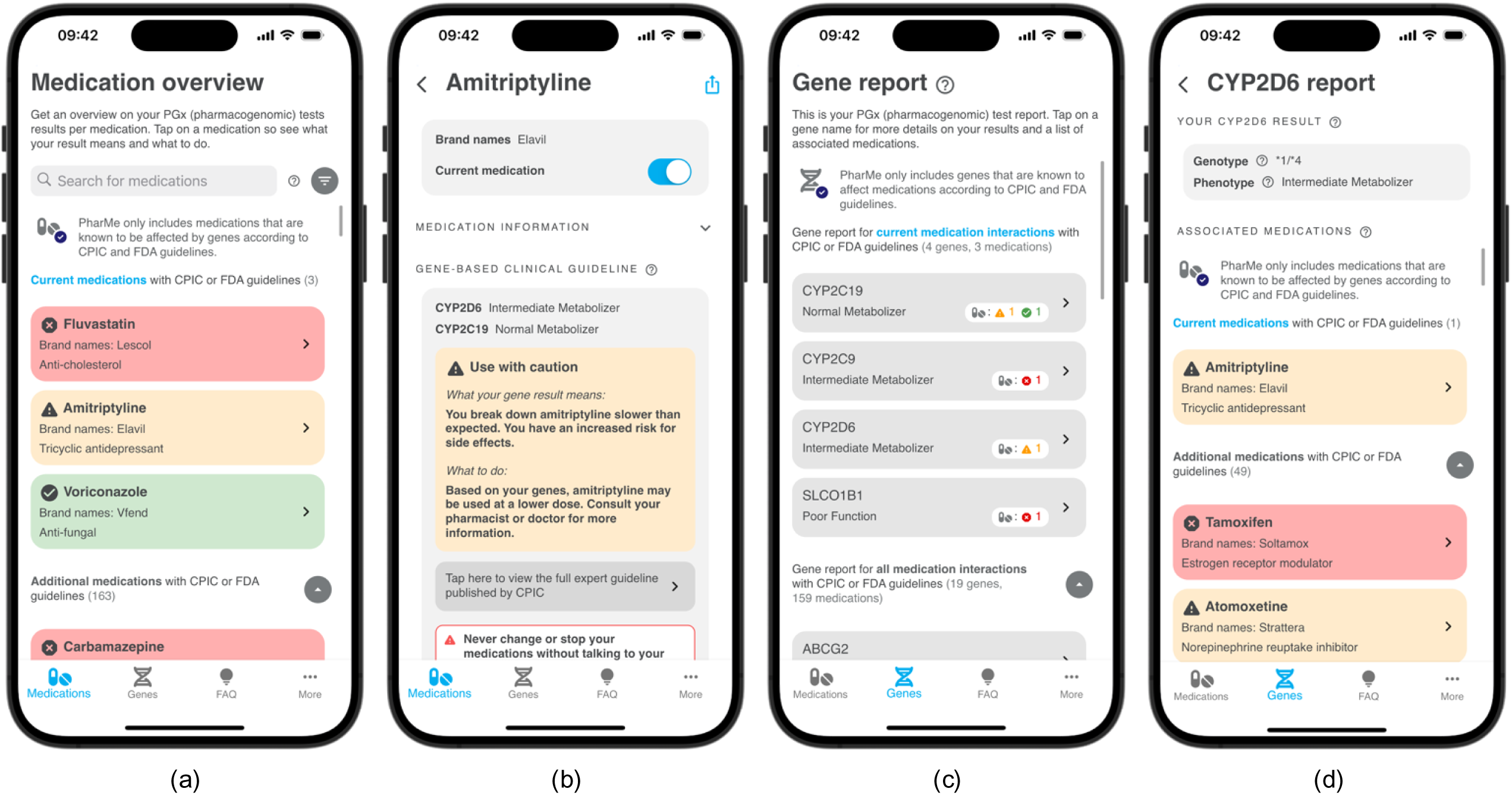
PharMe screens: (a) the Medication Overview screen with search and filter, (b) Medication screen containing annotated guideline information, (c) Gene Results screen, (d) Gene screen with implicated drugs.

**Figure 4.**
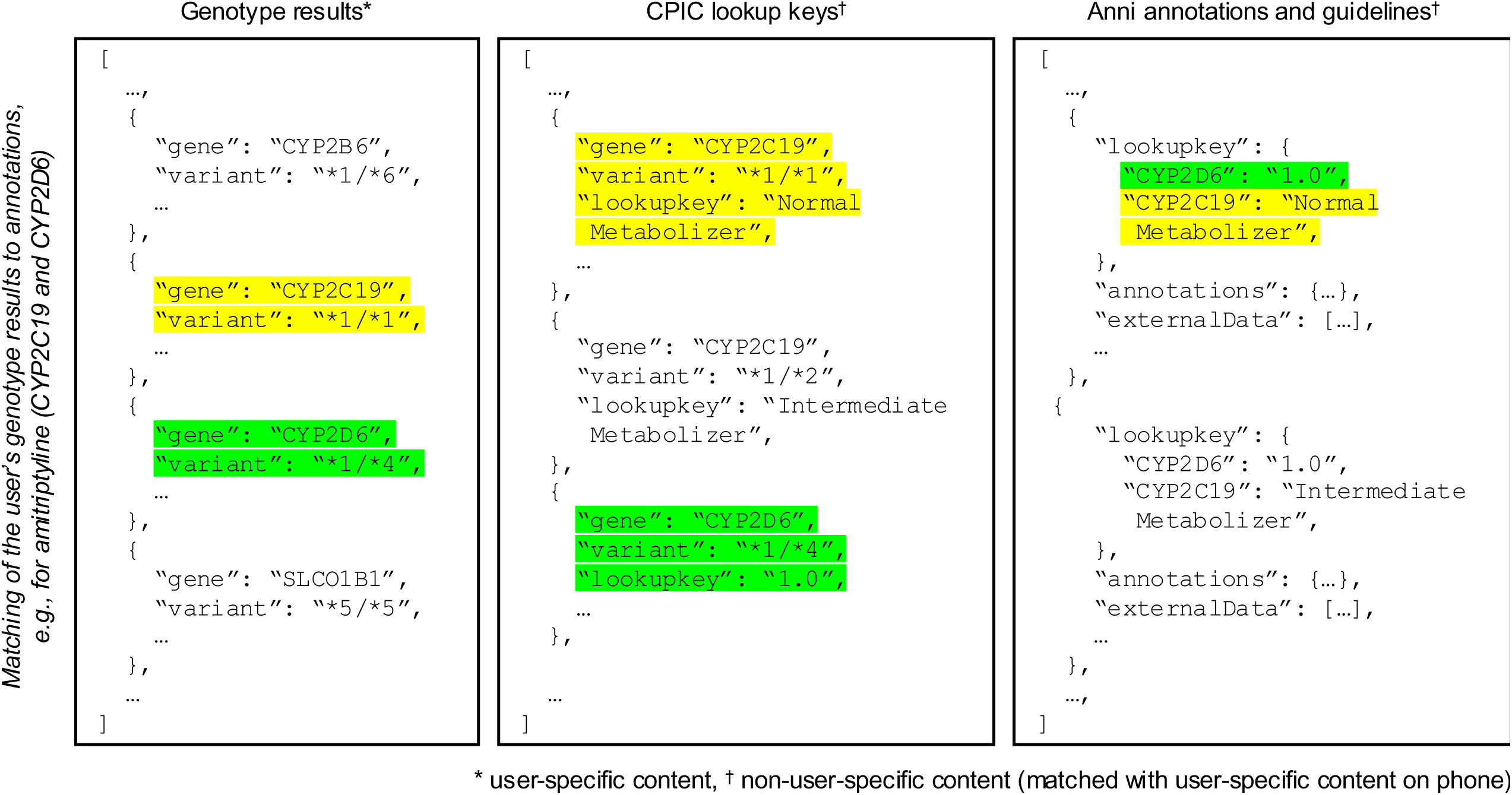
Format of data used in PharMe and mapping of PGx test results, guidelines, and annotations based on CPIC lookup keys.

After the initial login, users can update the list of imported active (or current) medications and review a short tutorial. The user is then directed to the Medication Overview screen (see Figure 3a). All medications included in PharMe are displayed in a color-coded list according to their respective guideline annotation warning level, with the user’s active medications listed first. In addition to the color-coding, icons are used to indicate the warning level to increase accessibility. A " None " warning level is used when no recommendation can be made because of missing genetic data or guidelines. This is displayed in green with a question mark icon to differentiate it from “green” warning levels. Users can search for medications by their generic name, brand names, or patient-friendly drug class descriptions, and filter the list by guideline warning levels. Users can tap on the medication for more information and the corresponding PGx guideline (see Figure 3b). Here, they can select whether they are currently taking the medication, export a PDF copy of their genetic results and the corresponding guideline recommendations to their healthcare provider, or access the references for the PGx-guided recommendations. We intentionally did not include a feature to show alternative medications or exact dose adjustments to discourage users from changing their prescribed medications and/or doses without consulting a healthcare professional. On the Gene Report screen, users can review their PGx test report per gene (see Figure 3c). Similar to the Medication Overview screen, genes associated with the users’ current medications are displayed at the top of the list. Tapping on a gene will lead users to the Gene screen (see Figure 3d), showing genotype and phenotype information and associated medications. Furthermore, PharMe includes tooltips and a frequently asked questions (FAQs), to provide further guidance (see Supplementary Figure S2c). On the More screen (see Supplementary Figure S2d), users can contact the app team with questions or problems and find additional resources about the app, genetics, and pharmacogenomics in general.

Moreover, PharMe supports phenoconversion (see Figure 5), currently implemented for moderate and strong CYP2D6 inhibitors. This functionality can be extended to cover additional drug-gene interactions in the future. When an inhibiting medication is selected as an active medication for the user, they will be presented with a warning dialog explaining that this medication can interact with other medications (see Figure 5a). The phenotype of the impacted gene (i.e., CYP2D6 phenotype changed to poor metabolizer) will be changed throughout the app if a strong inhibitor is selected (Figure 5b, Figure 5c, Figure 5d). If a moderate inhibitor is selected, the user’s phenotype is not changed; however, an interaction warning is displayed.

**Figure 5.**
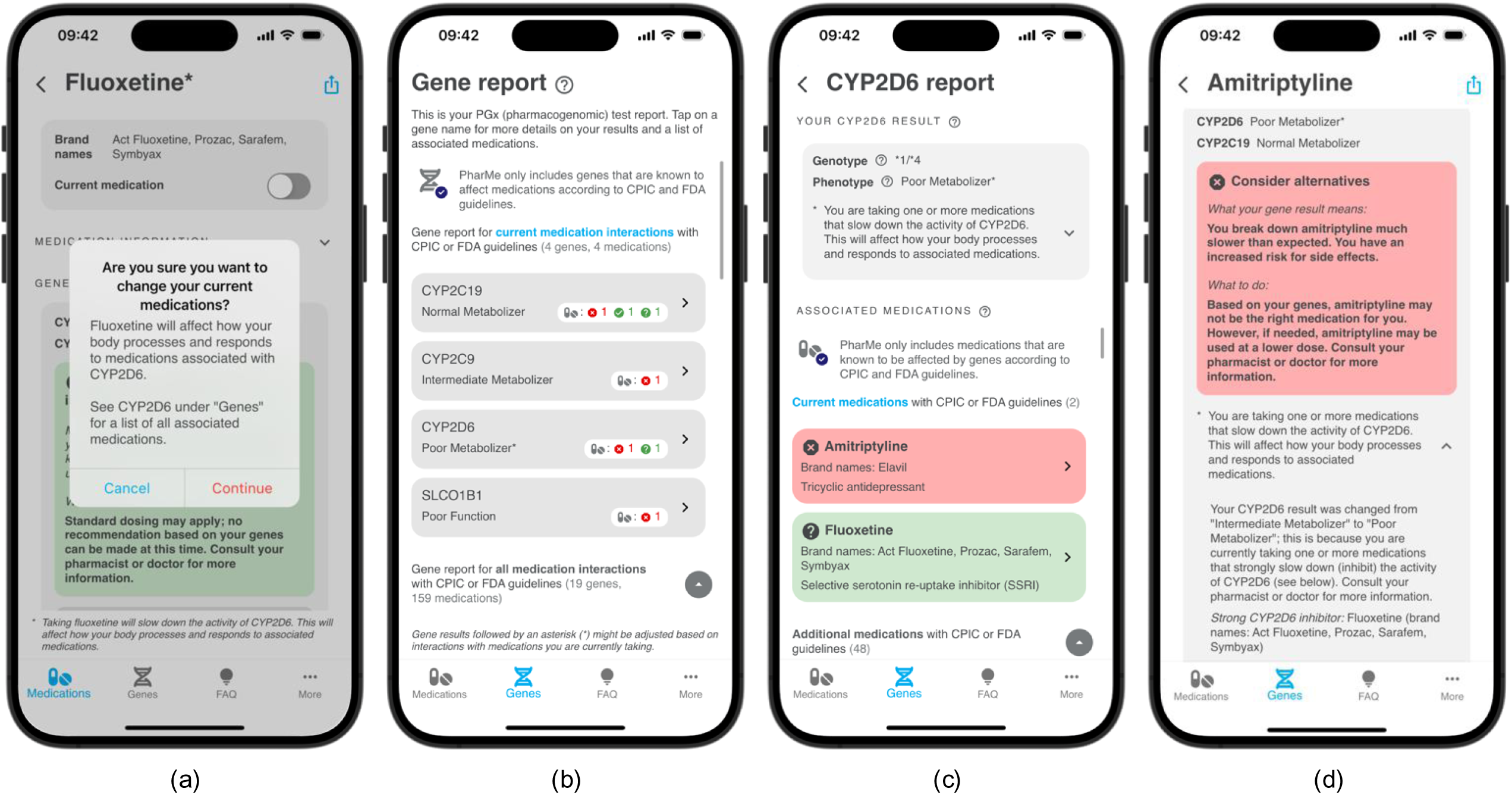
Phenoconversion in PharMe; (a) if a drug that is an inhibitor is selected as a current medication, a warning is shown; (b) the adapted phenotype is shown on the Gene Report screen, (c) on the Gene screen, and (d) on the Medication screen.

Since PharMe is intended to be used by patients rather than healthcare professionals, we aimed to clearly state its limitations and disclaimers throughout the app. For instance, notices reminding users that (1) medications and/or doses should never be changed without consulting a healthcare professional (e.g., Figure 3b); (2) PGx is only one important aspect when choosing the right medication and the right dose (e.g., Supplementary Figure S2a, Supplementary Figure S2c); and (3) PharMe only includes information about medications and genes that are included in clinical guidelines (e.g., Figure 3a, Figure 3c) are clearly displayed throughout the app. PharMe is currently available for iOS and Android in English (the authors can grant test access upon request), but multi-language support is feasible.

#### Exemplary Patient Health Data Source

For the PharMe development, an exemplary patient health data source was set up. The exemplary data server can be reached via a REST API built using Nest.js [43]. The server uses a Keycloak [44] instance for credential management and secure authentication, and a MinIO [45] instance for high-performance object storage, containing patient information in encrypted form. Both tools include web interfaces for credential and data management for convenient access. Additionally, a PostgreSQL [46] database stores a mapping of each Keycloak user to their respective health data in the MinIO object storage. All components can be easily deployed using Docker [47]. While the exemplary server is currently only used for development and an ongoing validation study, it demonstrates how a secure data infrastructure can be established.

#### Scio

The initial prototype included a self-developed module called Scio (Latin for “I know”)^††^ to assess users’ comprehension. The Flutter package aims to measure user comprehension of mobile applications based on custom questions, including multiple-choice, single-choice, or text answers. The questions can contain app context, e.g., in PharMe’s case, the user’s phenotype. User responses are collected anonymously on a Supabase [48] backend and can be used to refine or personalize the app content. The present PharMe version does not include Scio, as an ongoing validation study contains comprehension surveys on its own, outside of PharMe.

### User Validation Results

#### World Café

Four group testing sessions with 3-4 persons each were held on March 26, 2024, with a diverse set of 11 persons (20 to 72 years of age; students, employees, and pensioners). Overall, testers agreed that the app is handy and that having PGx information at hand would be beneficial. They liked (1) the color-coding (red, yellow, green), (2) that medications with “more severe” guideline results are shown first, and (3) that sharing guideline information with healthcare professionals does not require sharing the more sensitive genetic data. However, testers pointed out concerns about seeing a medication they are currently taking being displayed in red. They complained that this can deter users from taking those medications out of fear. Furthermore, testers found the gene names challenging and intimidating, so they suggested showing the medication-related information first, since that contains actionable information. Testers commented that the limitations of the app should be clearly defined so that users can understand what the tool does and cannot do, i.e., the app does not consider non-PGx factors. The difference between “green” guidelines with the checkmark symbol (“standard precautions”) and the question mark symbol (“standard precautions (incomplete data)”) was not entirely clear when only considering the screen showing all medications. However, the testers agreed that it is essential to differentiate between the two and that the green color-coding is more fitting than, e.g., coloring incomplete data grey, as both refer to standard precautions; also, when reading the description on the Medication screen, the meaning became clearer. Testers pointed out several further possible improvements, among smaller ones that the genes could be sorted by the highest warning level severity as well, a chat bot to ask questions about results and genetics, and that PharMe could be extended to include more comprehensible versions of package inserts, drug-drug interactions, and warnings if in acute cases current medications are taken longer than recommended.

#### Remote Pilot Testing

Twenty-eight persons participated in the remote pilot testing of the final prototype (**Error! Reference source not found.**, Supplementary Material S4) from April 29^th^ to June 29^th^, 2024.

**Table 1.**
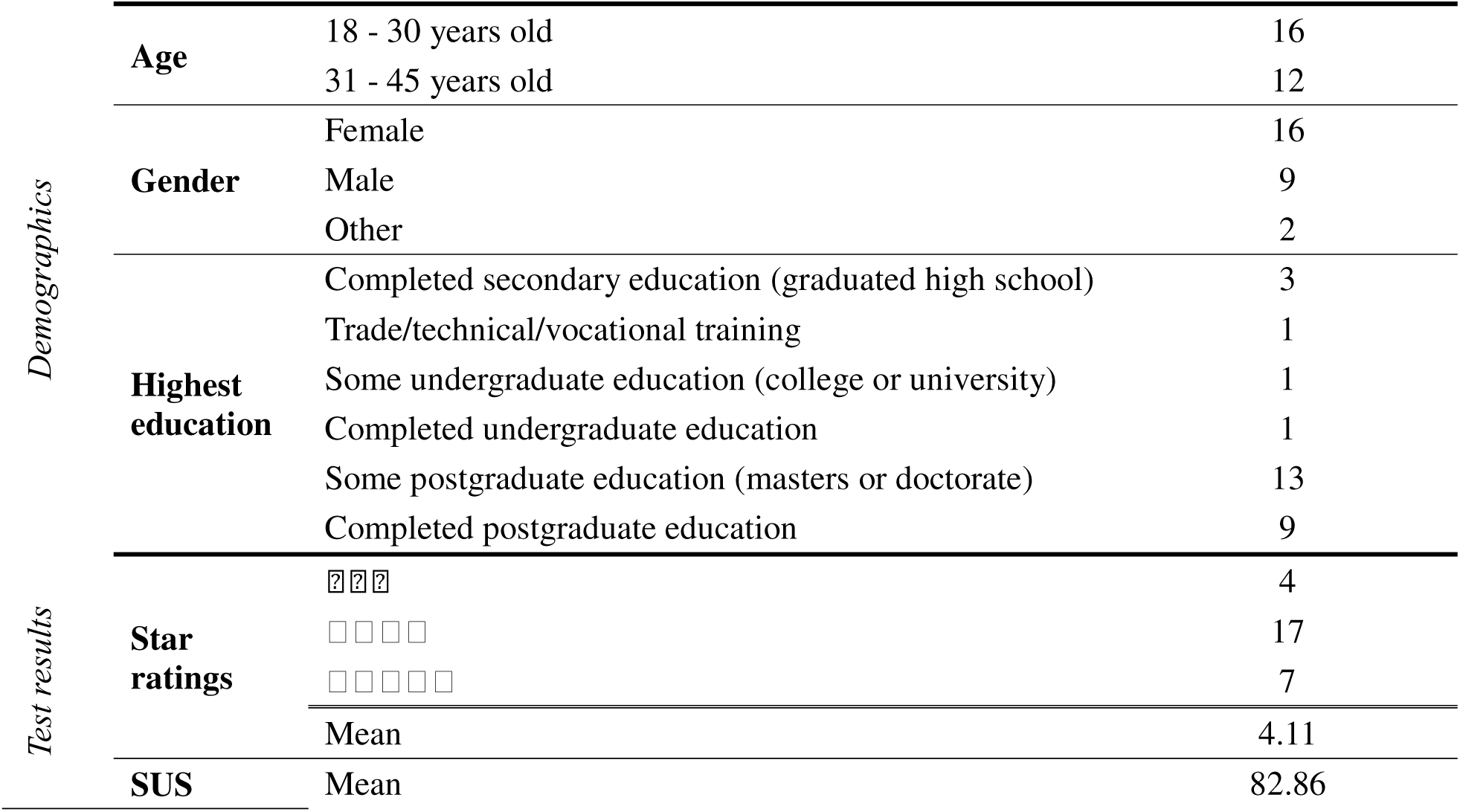
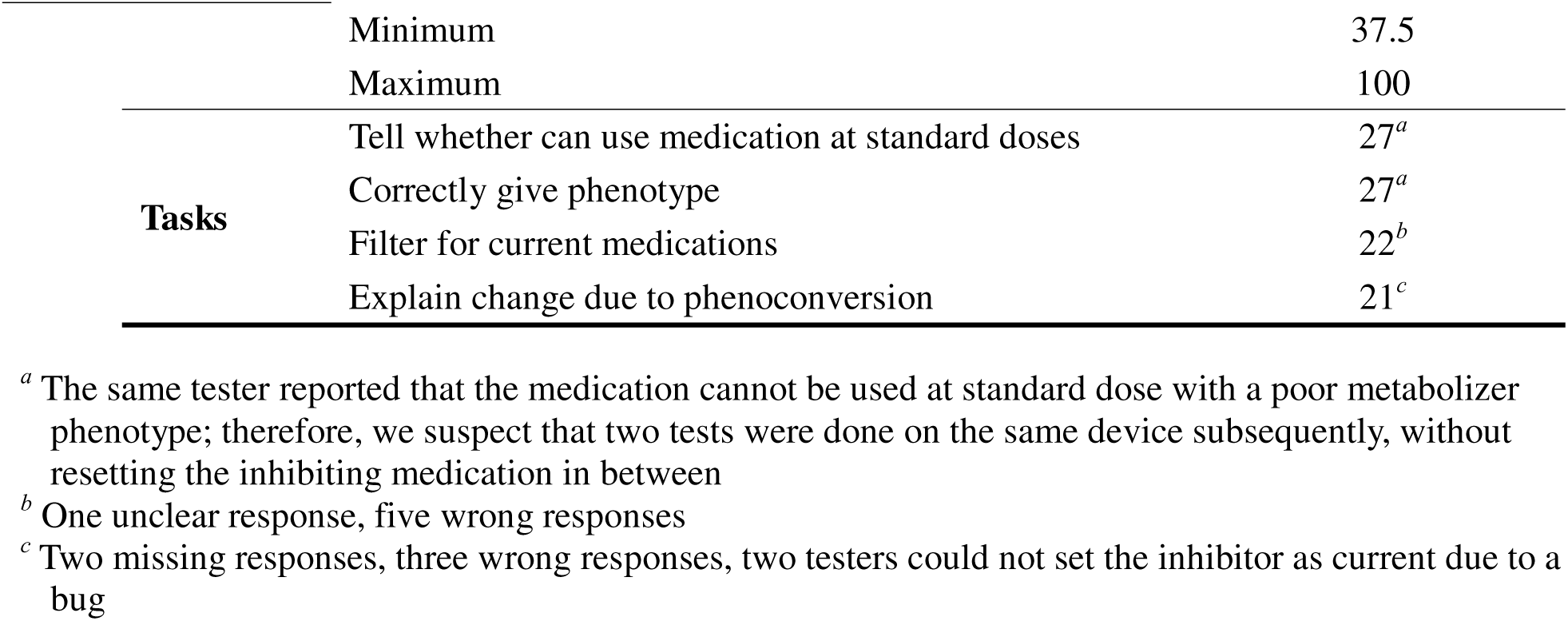
PharMe remote pilot testing results (total participants: 28).

|  |  |  |  |
| --- | --- | --- | --- |
| Demographics | Age | 18 - 30 years old | 16 |
|  |  | 31 - 45 years old | 12 |
|  | Gender | Female | 16 |
|  |  | Male | 9 |
|  |  | Other | 2 |
|  | Highest education | Completed secondary education (graduated high school) | 3 |
|  |  | Trade/technical/vocational training | 1 |
|  |  | Some undergraduate education (college or university) | 1 |
|  |  | Completed undergraduate education | 1 |
|  |  | Some postgraduate education (masters or doctorate) | 13 |
|  |  | Completed postgraduate education | 9 |
| Test results | Star ratings | ☆☆☆ | 4 |
|  |  | ☆☆☆☆ | 17 |
|  |  | ☆☆☆☆☆ | 7 |
|  |  | Mean | 4.11 |
|  | SUS | Mean | 82.86 |

|  |  |  |
| --- | --- | --- |
|  | Minimum | 37.5 |
|  | Maximum | 100 |
| **Tasks** | Tell whether can use medication at standard doses | 27<sup>a</sup> |
|  | Correctly give phenotype | 27<sup>a</sup> |
|  | Filter for current medications | 22<sup>b</sup> |
|  | Explain change due to phenoconversion | 21<sup>c</sup> |

<sup>a</sup> The same tester reported that the medication cannot be used at standard dose with a poor metabolizer phenotype; therefore, we suspect that two tests were done on the same device subsequently, without resetting the inhibiting medication in between
<sup>b</sup> One unclear response, five wrong responses
<sup>c</sup> Two missing responses, three wrong responses, two testers could not set the inhibitor as current due to a bug

The testers were younger, between 18 and 45, and had a high level of education. Most users were able to finish the tasks successfully. One person did not correctly identify that, according to the gene result, a medication can be used at standard dose and did not correctly select the phenotype; however, based on the tester’s selection, we suspect that two tests were done on the same device subsequently, without resetting the inhibiting medication in between. Six testers did not clearly explain how it is possible only to show results for current medications, and seven users could not explain phenoconversion, 2 of them because they could not activate the inhibiting medication due to a bug in the app. On average, PharMe achieved a star rating of 4.1 and a SUS score of 82.86, mapping to an “A” on the Sauro–Lewis CGS. The good ratings are in line with the textual feedback: overall, testers claimed that the app functions as intended, is well designed, and mostly runs stably, apart from bugs that were identified. However, the explanations given in PharMe could be more straightforward and comprehensible. One tester said they would only use the app if it were available in German. After the remote pilot testing, the app was adapted to include a list of current medications before all included medications to avoid the filter step, as already pointed out in the World Café. Also, additional information was provided for phenoconversion.

#### Power User Testing

The power user tested PharMe from October 15^th^, 2024, to February 06^th^, 2025. The tester confirmed the finding from the remote pilot testing that explanations should be adapted; they suggested a reading level corresponding to high school grades seven to nine to account for lower literacy levels. Additionally, they emphasized that the app should include clear explanations of what it is capable of, i.e., that PGx is only one important aspect when choosing the right medication and the right dose, and that PharMe only includes information about medications and genes included in clinical guidelines. The app was adapted accordingly, including adding to all annotations in Anni that the implications and recommendations can only be made “based on your genes”, further fixes of bugs and display problems, navigation improvements, and usability features, such as a “show password” button and a gene report for current medications, analogous to the list of current medications. The tester was consulted again throughout the process to obtain their feedback on the adaptations and re-iterate.

## Discussion

With PharMe, we developed an open-source service that transparently returns PGx test results to patients and is agnostic to data sources. Anni supports experts in creating consistent and multi-lingual patient-friendly annotations based on published guidelines.

Anni and PharMe are designed to support the most clinically relevant drug-gene interactions, currently focusing on CPIC and FDA guidelines and including 17 genes and 159 medications. While we currently only use a single source per guideline to simplify first annotations (e.g., ignore the FDA guideline if there is a CPIC guideline), the data structures support multiple sources already. Also, PharmGKB [49,50] could be used as a single source to include guidelines, supporting guidelines that are not yet included, e.g., from the Dutch Pharmacogenetics Working Group. To support genes currently not included in the CPIC API and therefore in PharMe, custom lookup keys that map genotypes to guidelines could be defined in Anni.

Present solutions for returning PGx test results are either commercial DTC tests, for which concerns were raised regarding their quality and data privacy [51–53], or integrated with a specific clinical decision support system, as done by Lipschultz et al. [17] PharMe aims to be lab- and institution-agnostic and can be deployed in any system. It also aims to keep and process patient health data only on the phone to increase data security and privacy. Further, our system transparently relies on standardized published guidelines and keeps user information safe and in the hands of users. The administrative dashboard developed by Lipschultz et al. is very similar to Anni, providing patient-friendly, rich-text content and media. While this approach allows for flexible and more extensive user reports, with Anni, we aimed to create consistent and concise formulations that can easily be translated into multiple languages. However, extending plain-text bricks to include rich-text and media content in the future may be reasonable.

PharMe was developed using a user-centric approach to make the app valuable, understandable, and easy to use. The remote pilot testing results indicate that PharMe meets this objective. Test users reported high satisfaction with the app in the star rating and the SUS, but ratings may be more positive as the remote pilot testing was conducted with peers. Still, the textual feedback agrees with the positive ratings, and most tasks were successfully solved, which indicates that the app is also objectively usable. However, most testers were younger and had higher education; therefore, we assume our test group to be more (digitally) literate than the general population. While our power user pointed out various design flaws that will affect their age group, they are also digitally literate. Despite these limitations, we improved the app with the feedback collected in all user discussions and testing sessions, especially by adapting the textual content to lower reading levels. Simple language is essential when designing understandable PGx reports, as Olsen et al. found in a study assessing patient understanding of printed CYP2D6 reports.[54] Moreover, they emphasized that PGx reports should include relevant information for the patient personally and actionable information, such as medications that are affected by a gene, which PharMe already covers. In addition, PharMe is being validated on a larger scale and with a more diverse population in a clinical non-inferiority study at Mount Sinai, comparing results returned by the app versus pharmacist-led counseling (Mount Sinai IRB protocol # 22-01404).

In the future, Anni could include further features such as directly providing drug information and brand names to annotators and using ontology-based indications. Also, additional factors to genetics that are important to be considered as part of PGx guidelines could be defined, such as age, current diagnoses, and ancestry. Further, the user interface could be extended to include currently scripted functionalities, such as updating guidelines, rule-based brick usage validation, showing annotation versions, and defining phenoconversion relations. PharMe could also be extended by further features such as: getting star allele information from VCFs or MSC codes directly, which would expand its applicability to data sources; considering drug-drug interactions; searching medications by scanning the barcode; or including a medication history rather than only current medications. While we focused on PharMe as a mobile application, it could also be deployed as a web service to work better if patients view their results at home, which is also supported by Flutter. Finally, Scio could be applied in practice again to monitor user comprehension in a first step or even personalize content in Anni and PharMe in a next step.

The immediate next steps should be to provide PharMe in more languages, using the present localization infrastructure, and including additional PGx guidelines. Further, we are currently investigating the use of large language models to create initial annotations, which would only require the subsequent review step and considerably speed up adding or updating guidelines. Moreover, the interaction between patients, PharMe, and healthcare professionals should be investigated, as we mainly focused on how patients use the app. The current exports for healthcare professionals should be validated and extended, and they could even be provided with MSC QR codes. Using MSC codes for reading and giving information would further strengthen PharMe’s integration in present initiatives like U-PGx.

## Conclusion

With PharMe and Anni, we developed a novel and user-friendly system to return PGx results to patients and scale the clinical adoption of PGx globally to support the limited number of trained clinical PGx experts. The system is designed to use transparent and standardized content based on published guidelines and to be agnostic to PGx data sources, which is why it can be flexibly deployed in different settings and institutions. A clinical validation study to assess PharMe on a larger scale and with a more diverse population at Mount Sinai is in progress.

## Lay Summary

It is well-known that pharmacogenomics (PGx), or how the genome influences the processing of medications in the body, can optimize therapeutic outcomes while minimizing adverse events. However, many barriers hinder its widespread adoption, such as the lack of trained healthcare professionals to adopt PGx into routine practice. To alleviate this burden, we have developed the PharMe app to return PGx results to patients, which transparently conveys easy-to-understand content, is based on published guidelines, is agnostic to PGx data sources, and can therefore easily be used in different settings and institutions. PharMe is powered by Anni, a novel annotation interface intended to support creating patient-facing content based on published guidelines, while enhancing consistency by re-using formulations and multi-language support. This paper describes the user-centered development of PharMe and Anni and shows the first promising user evaluations. A clinical validation study to assess PharMe on a larger scale and with a more diverse population at Mount Sinai is in progress. In all, we have developed a tool that can scale the clinical adoption of PGx globally to support the limited number of trained clinical PGx experts.

## Data Availability

All data produced in the present work are contained in the manuscript or available upon reasonable request to the authors

## Acknowledgements

We want to thank Tiffany Morris, Vlad Sima, and Volker Liebenberg for their support as corporate partners from Illumina in the student project that led to the initial prototype development. Further, we want to thank Jonathan A. Edelman, Felix Rebitschek, and all other experts and users who provided app feedback and valuable insights. We wish to express our sincere gratitude to Michelle Hou and Syeda Haque who helped to create the first version of guideline annotations as PharmD students, Sina Rampe, who significantly supported us to in the process to generate guideline annotations in Anni, and Matteo Danieletto and Kyle Landell, who provided substantial help with deploying PharMe on Mount Sinai infrastructure and its publishing in app stores, and Anna Stern, who invested considerable time and effort in testing PharMe through its paces.

## Author Contributions

T.S. and A.O.O. wrote the manuscript. E.P.B., C.L., S.M.M., J.B., B.F., N.G., L.H., S.K., C.M., M.S., and S.B. reviewed and extended the manuscript. J.B., B.F., N.G., L.H., S.K., C.M., M.S., S.B., and T.S. developed the software described in this manuscript. J.B., B.F., N.G., L.H., S.K., C.M., M.S., and S.B. created the PretoVid and conducted user interviews and testing leading to the initial prototype. T.S. conducted and evaluated further user and expert testing sessions.

## Supplementary Material

*Supplementary Figure S1. Additional Anni screens: (a) home page with publish button, (b) overview of annotations, (c) medication annotations and available guidelines per phenotype; (d) full annotations for amitriptyline (not visible in Figure 2); (e) overview of text bricks; (f) text brick details with German translation.*

*Supplementary Figure S2. Additional PharMe screens: (a) first onboarding screen, (b) login with patient data source selection, (c) FAQ screen, (d) More screen with further settings and information.*

*Supplementary Material S1. Interview guides used in the initial user and expert interviews.*

*Supplementary Material S2. Scripts used in World Cafe discussions.*

*Supplementary Material S3. Survey of the remote pilot testing.*

*Supplementary Material S4. Pilot testing results in CSV format (free text answers were removed or mapped to binary values for PharMe tasks).*

*Supplementary Material S5. List of included genes and medications.*

*Supplementary Material S6. PDF export for healthcare providers with and without phenoconversion.*

## Footnotes

‡ https://www.youtube.com/watch?v=iBCYnf_5oxc

§ https://github.com/hpi-dhc/PharMe

** https://github.com/hpi-dhc/PharMe-Annotations

†† https://github.com/hpi-dhc/scio

