## Supplementary figures and images for "Design and Implementation of PharMe: A Mobile Application to Return Pharmacogenomic Test Results to Patients"

### Supplementary Figure 1

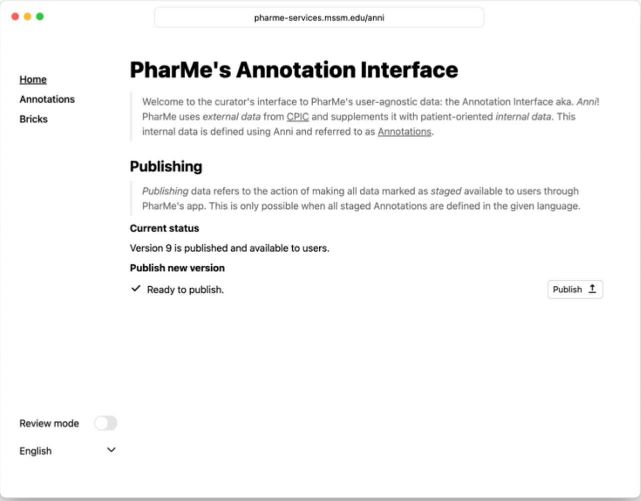

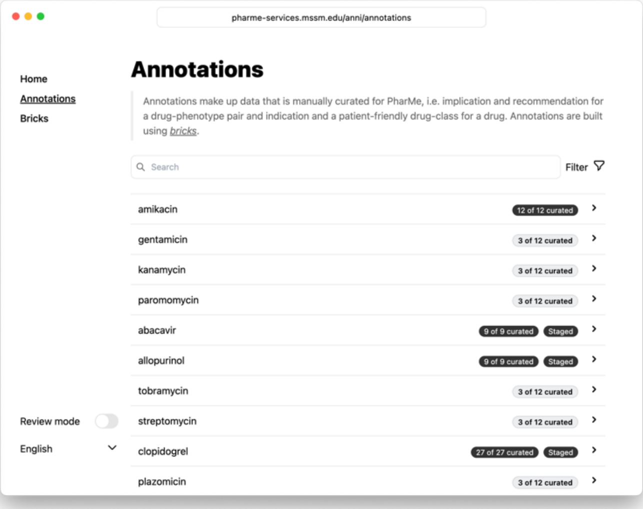

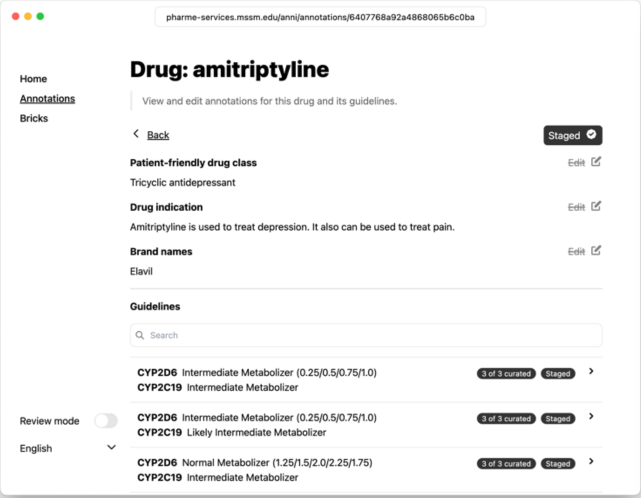

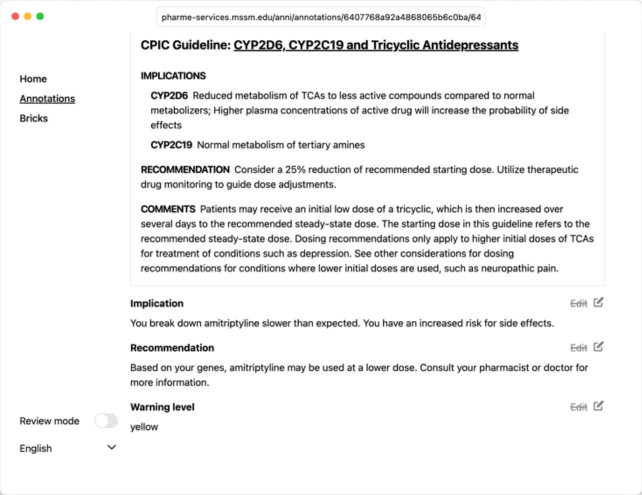

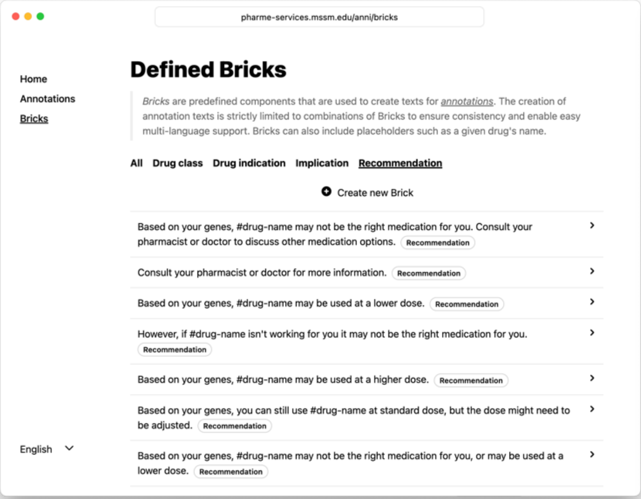

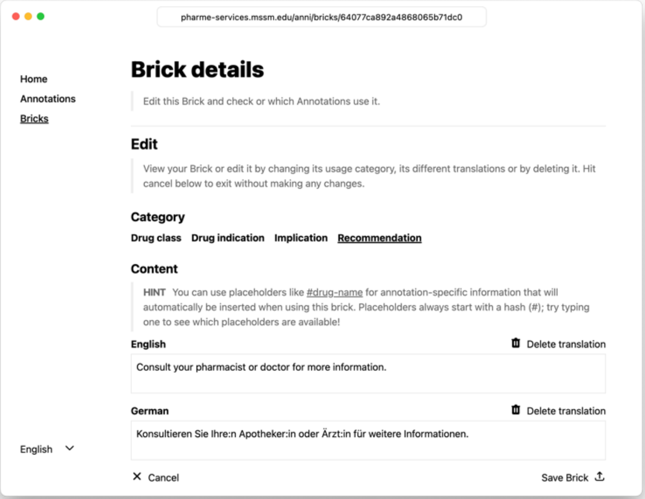

### Supplementary Figure 2

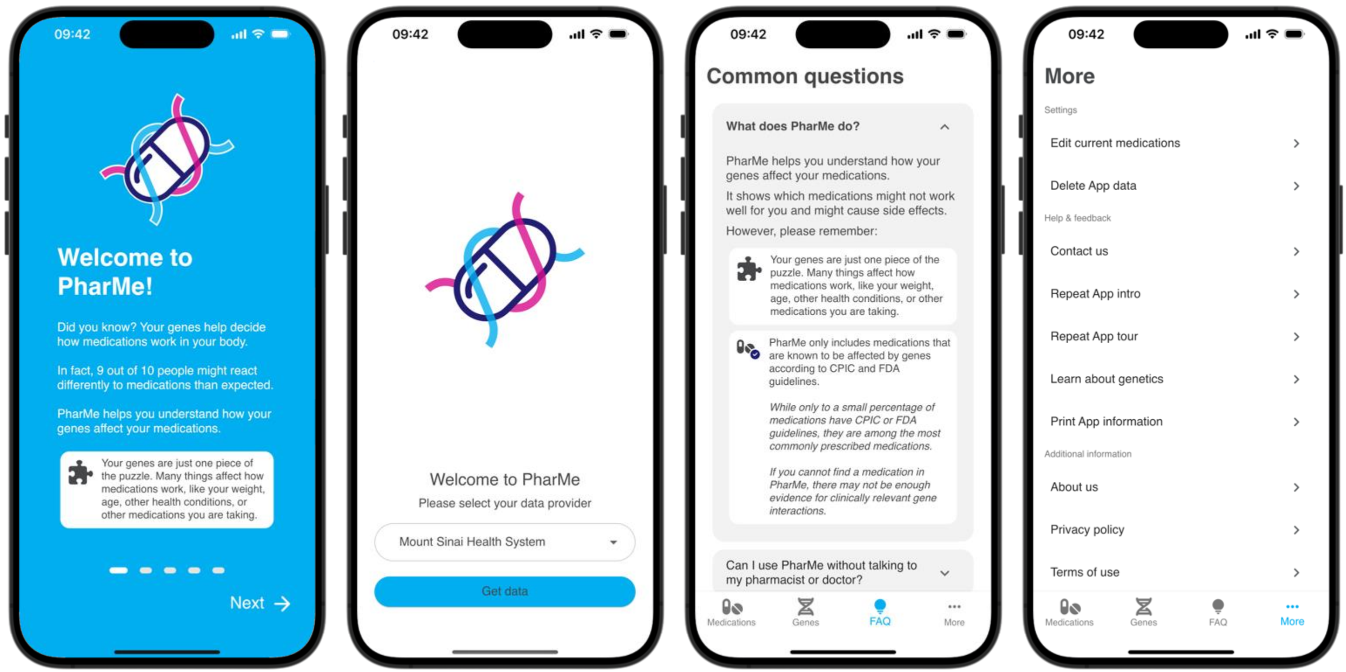
