## Supplementary Material 1 for "Design and Implementation of PharMe: A Mobile Application to Return Pharmacogenomic Test Results to Patients"

### PretoVid Interview (Users)

**Contents of the Interview**

Today, we will be conducting an interview that will be aimed at evaluating a use-case scenario in the form of a so-called preto video.

**Collection, Usage and Maintenance of Data**

Answers will be saved in the form of notes. No personal data [1] is being collected. Should you agree to this, the Interviewer keeps a random ID that was assigned to your name. This serves solely the purpose of being able to approach you with new questions at a later time. All collected data will be dedicated to improving the design and user experience of the app and to thereby improving the understanding of the app from the perspective of our target group. Collected data will be saved digitally.

**Rights regarding data protection**

The interviewee has the right to ask for deletion of his personal health information. However, data that was de-identified and aggregated can not be linked to the interviewee anymore and therefore, removed.

**Statement of Agreement**

I confirm that I understand the provided information, aims, procedures and risks of this interview. I have had the opportunity to ask questions, which were all answered satisfactorily. I understand and consent that my participation and the collection of my health data is voluntary. I understand that I can withdraw my consent at any time, without giving a reason and without any costs. I confirm that I received a copy of this consent form and interview description.

____________________________________________

**Date, Signature**

[1] https://ec.europa.eu/info/law/law-topic/data-protection/reform/what-personal-data_en

**Preparation: Data Protection Statement**

Hand out a version of the data privacy statement to the interviewee and explain the contents briefly. Answer any questions. Let them sign your own copy of the statement.

#### 1. Introduction and general questions

Thank you for taking the time to answer some of our questions! Not only are you helping us, but you are also helping to advance medicine.
We are developing a service as a part of our bachelor's thesis. Therefore - to better understand the point of view of the user - we would like to find out a bit about your relation to our topic. We will focus on that topic in a bit, don't worry.

Just some general questions:

a. How old are you?
b. What's your occupation?
c. What is the highest degree or level of school you have completed?
d. Do you own a smartphone?

#### 2. Preto Video

We have put together a video that briefly presents the medical background, our service and our goal. Now we would like to test how comprehensible we have made the contents of the video to improve upon it and learn from it.

#### 3. Comprehension: Case Study and Idea of the App

***Show video (0:00 to 1:33)*** ([https://www.youtube.com/watch?v=iBCYnf_5oxc](https://www.youtube.com/watch?v=iBCYnf_5oxc))

a. How did the case study without PharMe transpire in your understanding?
b. Were there aspects of this case study that made it hard for you to identify with the patient? (e.g. was something unrealistic?) If so, what?
c. How did the case study with PharMe transpire in your understanding?
d. Which of the two case studies did you find more desirable? Which aspects of the video have motivated you for/against PharMe?

Previously, you have (not) mentioned data privacy as a reason for/against PharMe. In this next section we want to touch upon this topic in detail.

**4. Comprehension: Data Protection**

a. How did you perceive the data flow of sensible data from the video? Where will your data be stored?
b. Were there aspects of the video that positively or negatively influenced your considerations regarding data protection?

***If the interviewee did not completely understand the part of the video dedicated to data privacy, please now respectfully clarify them, so the next answer is qualified***

c. Should there be more focus on data protection in the video? If so, what could be communicated better?

**5. Comprehension: Pharmacogenomics/Pharmacogenetics (PGx)**

***Show video (1:33 to 2:50)**** (https://youtu.be/iBCYnf_5oxc?t=93)

Those were the most important medical cornerstones of our service. We would also like to test the understandability of this part of the video.

a. How affected do you feel by the field of research that was being discussed? Does it seem relevant to you?
b. [If interviewee feels affected] Which parts of the video have contributed to this?
   [If interviewee does not feel affected] Why do you not feel affected by this field of research?
c. Which part of this section did you find most impressive?

***Show video (2:50 to end)**** (https://youtu.be/iBCYnf_5oxc?t=170)

**6. Concluding Questions**

First and foremost we want to thank you for taking the time to answer our questions. For what it is worth you really helped us. And not only that: Our service is going to be used for research in the field of Pharmacogenomics, aiming to provide patients and their care providers with the latest research in order to support informed decision making. Having taken the time for this interview, you are helping to advance the field of personalized medicine.
If you have any other suggestions regarding the video or our service please let us know.
***note down feedback***

To conclude, if there was anything you have noticed about the conduct of the interview or the questions, we would also be happy about feedback in this context. ***note down feedback***

### Expert Interview (Medical Professional)

**Contents of the Interview**

Today, we will be conducting an interview that will be aimed at evaluating the mockups of an app and a use-case scenario in the form of a so-called preto video. The second section will consist of questions about the participant and the underlying domain.

**Collection, Usage and Maintenance of Data**

Answers will be saved in the form of notes. No personal data [1] is being collected. Should you agree to this, the Interviewer keeps a random ID that was assigned to your name. This serves solely the purpose of being able to approach you with new questions at a later time. All collected data will be dedicated to improving the design and user experience of the app and to thereby improving the understanding of the app from the perspective of our target group. Collected data will be saved digitally.

**Rights regarding data protection**

The interviewee has the right to ask for deletion of his personal health information. However, data that was de-identified and aggregated can not be linked to the interviewee anymore and therefore, removed.

**Statement of Agreement**

I confirm that I understand the provided information, aims, procedures and risks of this interview. I have had the opportunity to ask questions, which were all answered satisfactorily. I understand and consent that my participation and the collection of my health data is voluntary. I understand that I can withdraw my consent at any time, without giving a reason and without any costs. I confirm that I received a copy of this consent form and interview description.

________________________________________

**Date, Signature**

**Goals of the interview**

- **Are other tools currently used for helping decision making in practices/clinics?**
- **Determine conscience regarding PGx**
- **Would doctors use our app?**
- **What concerns are there?**
- **-> Preto Video**

**Preparation: Data Protection Statement**
[First question: German or English preferred?]

[Hand out a version of the data privacy statement to the interviewee and explain the contents briefly. Answer any questions. Let them sign your own copy of the statement.]

First of all, thank you so much for sparing some time for this interview. (As previously mentioned:) We are students at the HPI in Potsdam, currently working on our bachelor's in computer science. In the scope of a software project, we are developing a service for supporting patients and medical professionals and thus would like to understand the doctor's point of view better.

Let's start with a few basic questions:

1. A. What field are you specialized in?
   B. Are you established or working in a clinic?
   C. Since when?
2. A. When several paths of treatment are worth considering, how do you make your decision on which path to choose?
   B. Are any tools involved in this process? If so, which ones?
3. A. What role do interindividual differences play in efficacy and side effects of medications in your everyday life?
   B. What are the most important factors causing these differences?
4. A. Have you ever heard about pharmacogenomics? What do you know about the field?

We now want to present to you our service that is currently in development. For this we prepared a video. Since it is mainly for people without any medical expertise, the language is a bit simplified and you will most likely already be familiar with the scientific background. Nevertheless we would like to show you this video, since it presents the idea of our service quite concisely.
[Show them the entire preto video: https://www.youtube.com/watch?v=iBCYnf_5oxc]

5. A. To understand if our concepts were comprehensively depicted. How does the process between a potential user of the app in the form of a patient and their doctor play out according to your understanding of the video?
   B. Were there parts of this process that were unclear or not elaborated upon enough?
   C. How would you like to be presented with the results?
   D. Do you see a benefit worth the effort in this service? Why (not)?

E. If all regulatory conditions are fulfilled, could you imagine recommending this service to your patients? If so: How would you integrate the service in your decision making?
F. What other functions would be helpful?
G. Are there other scenarios where using our service would seem helpful?

6. Do you have any other thoughts on the service, the video or the general topic, which you would like to share?

First and foremost we want to thank you for taking the time to answer our questions. For what it is worth you really helped us. And not only that: Our service is going to be used for research in the field of Pharmacogenomics, aiming to support informed decision making. Can we contact you for further interviews about new, improved versions of our service and our ideas?

### Expert Interview (Digital Health Professional)

**Contents of the Interview**

Today, we will be conducting an interview that will be aimed at evaluating the mockups of an app and a use-case scenario in the form of a so-called preto video. The second section will consist of questions about the participant and the underlying domain.

**Collection, Usage and Maintenance of Data**

Answers will be saved in the form of notes. No personal data [1] is being collected. Should you agree to this, the Interviewer keeps a random ID that was assigned to your name. This serves solely the purpose of being able to approach you with new questions at a later time. All collected data will be dedicated to improving the design and user experience of the app and to thereby improving the understanding of the app from the perspective of our target group. Collected data will be saved digitally.

**Rights regarding data protection**

The interviewee has the right to ask for deletion of his personal health information. However, data that was de-identified and aggregated can not be linked to the interviewee anymore and therefore, removed.

**Statement of Agreement**

I confirm that I understand the provided information, aims, procedures and risks of this interview. I have had the opportunity to ask questions, which were all answered satisfactorily. I understand and consent that my participation and the collection of my health data is voluntary. I understand that I can withdraw my consent at any time, without giving a reason and without any costs. I confirm that I received a copy of this consent form and interview description.

________________________________________

**Date, Signature**

**Goals of the interview**

- **Find out opinions, concerns, experience about our idea: use case? concept? Approval? Financing?**
- **Would you use this service for yourself?**

**Preparation: Data Protection Statement**
[First question: German or English preferred?]

[Hand out a version of the data privacy statement to the interviewee and explain the contents briefly. Answer any questions. Let them sign your own copy of the statement.]

First of all, thank you so much for sparing some time for this interview. (As previously mentioned:) We are students at the HPI in Potsdam, currently working on our bachelor's in computer science. In the scope of a software project, we are developing a service for supporting patients and medical professionals and thus would like to understand the medical and digital-health point of view better.

1. First, a few basic questions:
   A. What specialty do you work in?
   B. Since when?
   C. What is your professional background?
   D. What are your main topics?

We would now like to introduce you to the service we are developing. For this purpose we have created a video. This was created primarily for people without medical or technical expertise, so the language is somewhat simplified and the background may already be familiar to you. Nevertheless, we would like to show you the video because it succinctly presents the idea behind our service.
[Show full Preto Vid: https://www.youtube.com/watch?v=iBCYnf_5oxc]

2. A. To understand if our presentation in the video was understandable, what did you remember about the steps needed from the doctor and the patient to use the service?
   B. Were there any parts of the process that were unclear?
   C. Do you see any added value in this service? Why (not)?
   D. What other functions could be helpful?
   E. If all regulatory conditions are met, would you use this service yourself?
   F. Would you recommend it to others?

3. Do you have any other thoughts on the topic, the video, or our service that you would like to express? (All directions possible, feel free to ask if needed).

First and foremost we want to thank you for taking the time to answer our questions. For what it is worth you really helped us. And not only that: Our service is going to be used for research in the field of Pharmacogenomics, aiming to provide patients and their care providers with the latest research in order to support informed decision making. Having

taken the time for this interview, you are helping to advance the field of personalized medicine.
Can we contact you for further interviews about new, improved versions of our service and our ideas?

### Wireframe Interview

**Contents of the Interview**

Today, we will be conducting an interview that will be aimed at evaluating the mockups of an app.

**Collection, Usage and Maintenance of Data**

Answers will be saved in the form of notes. No personal data [1] is being collected. Should you agree to this, the Interviewer keeps a random ID that was assigned to your name. This serves solely the purpose of being able to approach you with new questions at a later time. All collected data will be dedicated to improving the design and user experience of the app and to thereby improving the understanding of the app from the perspective of our target group. Collected data will be saved digitally and deleted after the project ends.

**Rights regarding data protection**

The interviewee has the right to ask for deletion of his personal health information. However, data that was de-identified and aggregated can not be linked to the interviewee anymore and therefore, removed.

**Statement of Agreement**

I confirm that I understand the provided information, aims, procedures and risks of this interview. I have had the opportunity to ask questions, which were all answered satisfactorily. I understand and consent that my participation and the collection of my health data is voluntary. I understand that I can withdraw my consent at any time, without giving a reason and without any costs. I confirm that I received a copy of this consent form and interview description.

____________________________________________

**Date, Signature**

**Preparation: Data Protection Statement**

Hand out a version of the data privacy statement to the interviewee and explain the contents briefly. Answer any questions. Let them sign your own copy of the statement.

**1. Introduction and general questions**

We are developing an app called PharMe as a part of our bachelor’s thesis. Therefore - to better understand the point of view of the user - we would like to find out a bit about your relation to our topic. We will focus on that topic in a bit, don’t worry.

For now we have some general questions:

- How old are you?
- What’s your occupation?
- What is the highest degree or level of education you have completed?
- Do you own a smartphone?
- Do you have a professional background in medicine?

To get an unskewed perspective, we intentionally will not explain the idea, purpose and usage of our product for now.

**2. Onboarding**

When you first download the app, you are presented with the following set of screens as an onboarding. In this first part of the interview let's just focus on that onboarding section. Please take your time and read it. [Stop the time and see how long users spend on this part]

[General question]:

- Would you entrust PharMe with your genomic data? If not, why wouldn't you trust PharMe?
- Would you continue exploring the app?
  - [If not] What are the reasons you wouldn't continue?
- After looking at the Onboarding would you consider taking a gene test to use all the apps advantages?

[Comprehensive questions]:

- Describe the problem PharMe intends to solve and the way PharMe intends to solve it in your own words.
- What benefit would PharMe deliver to YOU personally or to users in general according to your understanding?

[Design-related questions]:

- Did you find the onboarding process too long/informationally bloated?
- Colors
  - Did any color or gradient seem inconsistent with the contents of the screen or the illustration for you? If so, which ones?
  - [Show them the uncolored logo] If you had to choose a color/gradient for the corporate identity of PharMe?
  - (Do you have any other comments or feedback regarding colors in onboarding as well as branding?)
- Illustrations
  - *We are deferring the final design of these illustrations until later so we want to apologize for the eye pain for now*
  - Was there an illustration that seemed unclear to you? If so, which one and what did you think about it?
  - Are there illustrations that were contextually flawed? If so, which ones and what was flawed?
- (Do you have any other comments or feedback on the onboarding process in its entirety or any aspect about it?)

**3. Rest of prototype**

[For each task you assign to the user, stop the time and note down every time they interact with the app. After each of the following tasks, reset the app to the root route. (excluding OB)]

[Tasks]

- Find out if Amitriptyline would be suited for you!
- Find out why Amitriptyline is suitable or not!
- [if the interviewee didn’t already find it] Where would you find further information on why Amitriptyline is suitable or not?
- How would you share information with your doctor?
- Find out where PharMe gets its data!
- Delete your imported genomic data!
- Revisit the Onboarding pages!

[After finishing the tasks, ask the participant if he wishes to explore the prototype for a bit. When he is done, make the prototype inaccessible. The following questions should be answered from memory and not by repeating what’s written on the pages]

[Comprehensive questions]

- Would you use PharMe to select a proper medical treatment?
    - [if not] Why?
- What do you do after finding out that a medication is not suited for you?
- What does it mean to be an ultrarapid metabolizer?

[Questions specific to app segments]
[Show them the login process/login screens again]

- Where do you get the login credentials you need to type in the app?

[Show them the details screen]

- Was it easy for you to quickly find out if a specific medication is suited for you?
- Did you find the amount of information on the screen overwhelming?
    - [if so] What would you not include in this screen?
- Was it clear to you, how to find further information?

[Show them the reports page]

- Is the design easily understandable?
- [Without checking] What kind of medications are listed under the “Dose”-tab?
- Would you prefer all medications with warnings to be in one list or separated in these tabs? If one list, should they be grouped by ‘severity’? Sorted by what?

[Show them the PGx page]

- Look at the Questions and their answers! Are they easy to understand and comprehensive?
- Are there any questions that should also be answered here? Or are there unnecessary questions?

- (Do you have any other comments or feedback on the app in its entirety or any aspect about it?)

**4. Other/Concluding Questions**

First and foremost I want to thank you for taking the time to answer our questions. For what it is worth you really helped us.
If you have any other suggestions regarding the app or our service in general please let me know. ***note down feedback***

To conclude, if there was anything you have noticed about the conduct of the interview or the questions, we would also be happy about feedback in this context. ***note down feedback***
