## Supplementary Material 2 for "Design and Implementation of PharMe: A Mobile Application to Return Pharmacogenomic Test Results to Patients"

*[Translated from the original German version by author]*

### PharMe Testing Script (on iPad)

*The aim is to get to point 5; if the time in a group is up, jump to point 8 as last question.*

0. **Preparation**:
   - Testers have already seen the onboarding and tutorial as an introductory presentation
   - Active medications: Pantoprazole, Amitriptyline, Mimpara
   - (maybe deactivate Buprion after point 5)
   - Go to report page
1. **Introduction**:
   - I would like to test some PharMe functions with you; to do so, I prepared the app on an iPad.
   - I explain the task and you show me, how you would interact with the App to solve it.
   - **How to give feedback**:
     - In case you have questions or remarks that you would like to add to the feedback grid[1], please write them on a post-it, we will talk about your feedback after each task.
     - Please write the task number in one corner of your post-it (I will tell you which is the current task).
   - **Onboarding & Tutorial**: Before we start, do you have any feedback or questions regarding the introductory presentation I showed earlier?
2. **Report Screen**:
   - Post-it number: 2
   - Please have a look at the Report Screen
   - Do you have any feedback for this screen? What does work, what does not work, what is unclear?
   - How would you select **CYP2D6**? (*Tap on CYP2D6*)
3. **Gene Page**
   - Post-it number: 3
   - Have a look at the gene page
   - Do you have any feedback for this screen? What does work, what does not work, what is unclear?
4. **Medications Navigation**:
   - Post-it number: 4
   - Navigate to the Medications screen

[1] https://www.ibm.com/design/thinking/page/toolkit/activity/feedback-grid

- How would you show all medications that you are currently taking? (*Filter* > *“Current medications”*)
- Select **atomoxetine** (*need to remove filter!)*
- Do you have any feedback for this screen? What does work, what does not work, what is unclear?

5. **Medication Page**
   - Post-it number: 5
   - Based on the results, can you take atomoxetine in standard dose? (*Yes*)
   - Do you have any feedback for this screen? What does work, what does not work, what is unclear?
6. **Select a medication as active medication**
   - Post-it number: 6
   - How would you select **bupropion** as a medication that you are currently taking? (*either Medications* > *search/scroll (no “current medication” filter)* > *select drug* > *select as “Drug usage status” the option “I am currently using...”; or via More* > *“Current medications”*)
   - Do you have any feedback for this screen? What does work, what does not work, what is unclear?
7. **Back to Atomoxetine**
   - Post-it number: 7
   - Go back to Atomoxetine
   - Can you explain why the result changed? (*Bupropion inhibits CYP2D6*)
   - Do you have any feedback for this screen? What does work, what does not work, what is unclear?
8. **Star Rating**
   - Post-it number: 8
   - If you would give an App Store rating for PharMe, how many stars would you give (1–5)?
   - Please give a short explanation, why
9. **Are there further pages or functions that you would like to explore?** (*If yes: ask “default question”, Do you have any feedback for this screen? What does work, what does not work, what is unclear?*)
10. **Further topics for discussion**
    - What is unclear? Do you have any open questions? *(Please add them to questions as post-its.)*
    - Would you like to use PharMe? Why (not)?
    - What would be concerns if you were using PharMe?

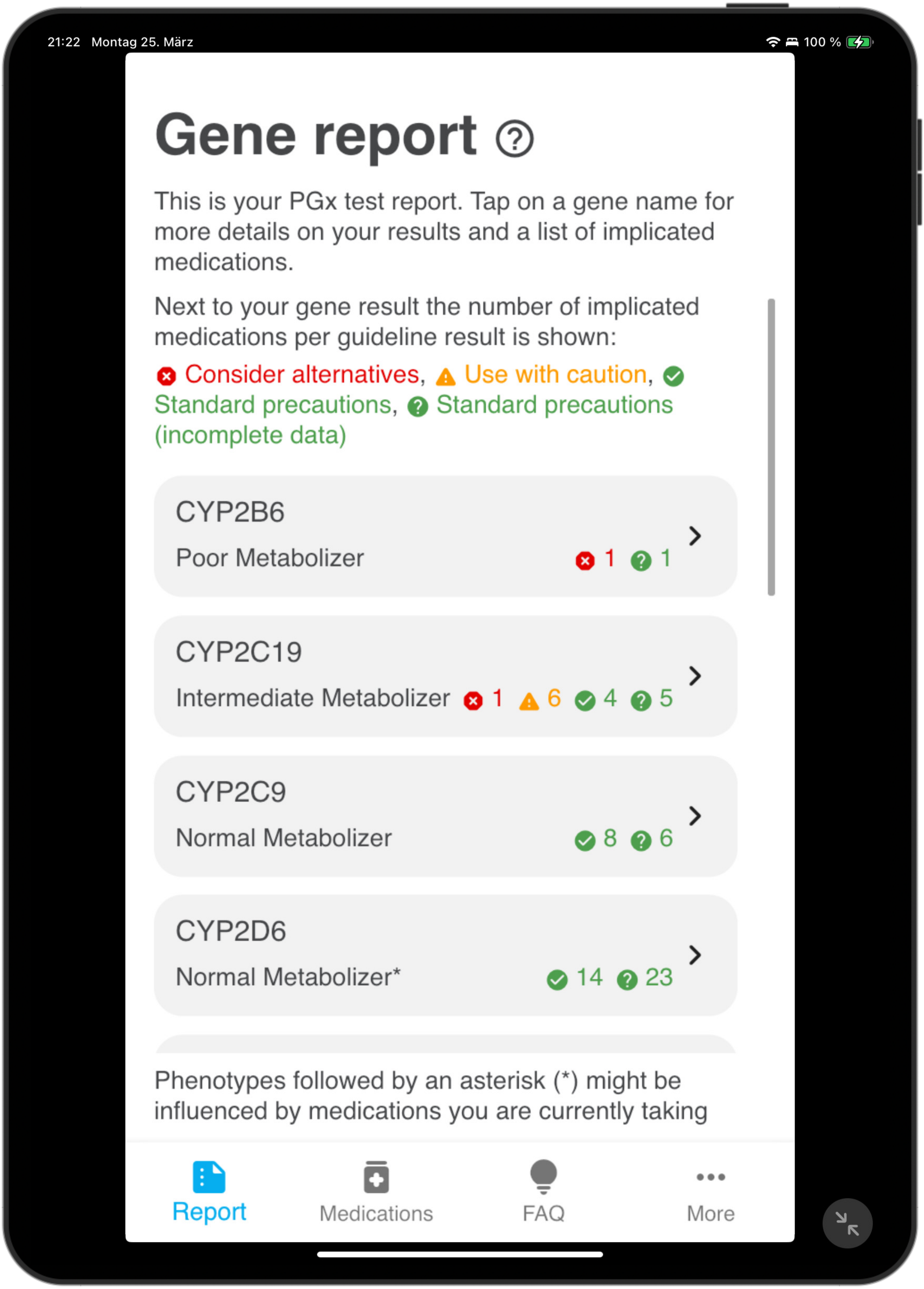
21:22 Montag 25. März
100 %
Gene report
This is your PGx test report. Tap on a gene name for more details on your results and a list of implicated medications.
Next to your gene result the number of implicated medications per guideline result is shown:
Consider alternatives, Use with caution, Standard precautions, Standard precautions (incomplete data)
CYP2B6
Poor Metabolizer 1 1
CYP2C19
Intermediate Metabolizer 1 6 4 5
CYP2C9
Normal Metabolizer 8 6
CYP2D6
Normal Metabolizer* 14 23
Phenotypes followed by an asterisk (*) might be influenced by medications you are currently taking
Report
Medications
FAQ
More

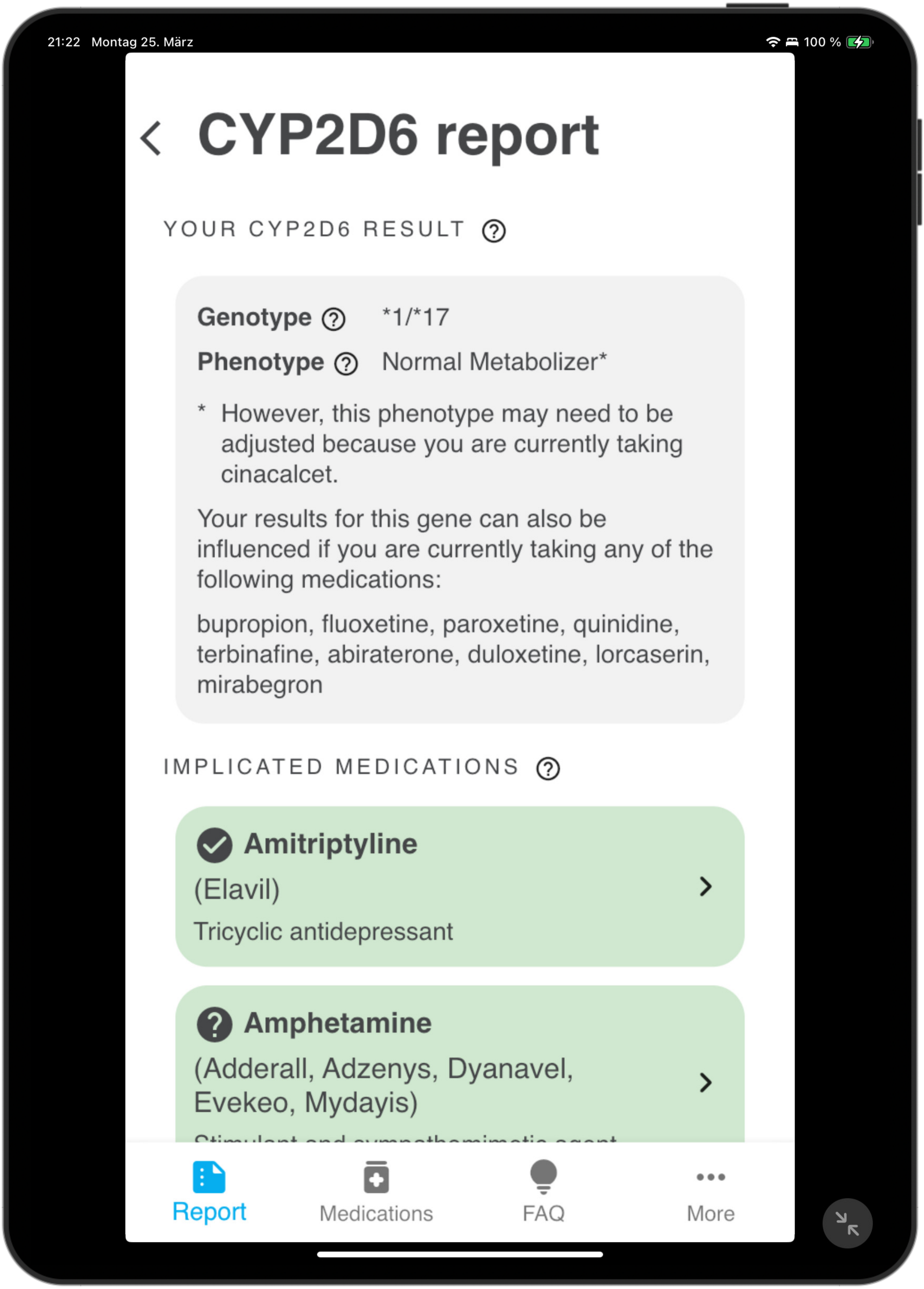
21:22 Montag 25. März
100 %
CYP2D6 report
YOUR CYP2D6 RESULT
Genotype *1/*17
Phenotype Normal Metabolizer*
* However, this phenotype may need to be adjusted because you are currently taking cinacalcet.
Your results for this gene can also be influenced if you are currently taking any of the following medications:
bupropion, fluoxetine, paroxetine, quinidine, terbinafine, abiraterone, duloxetine, lorcaserin, mirabegron
IMPLICATED MEDICATIONS
Amitriptyline
(Elavil)
Tricyclic antidepressant
Amphetamine
(Adderall, Adzenys, Dyanavel, Evekeo, Mydayis)
Report
Medications
FAQ
More

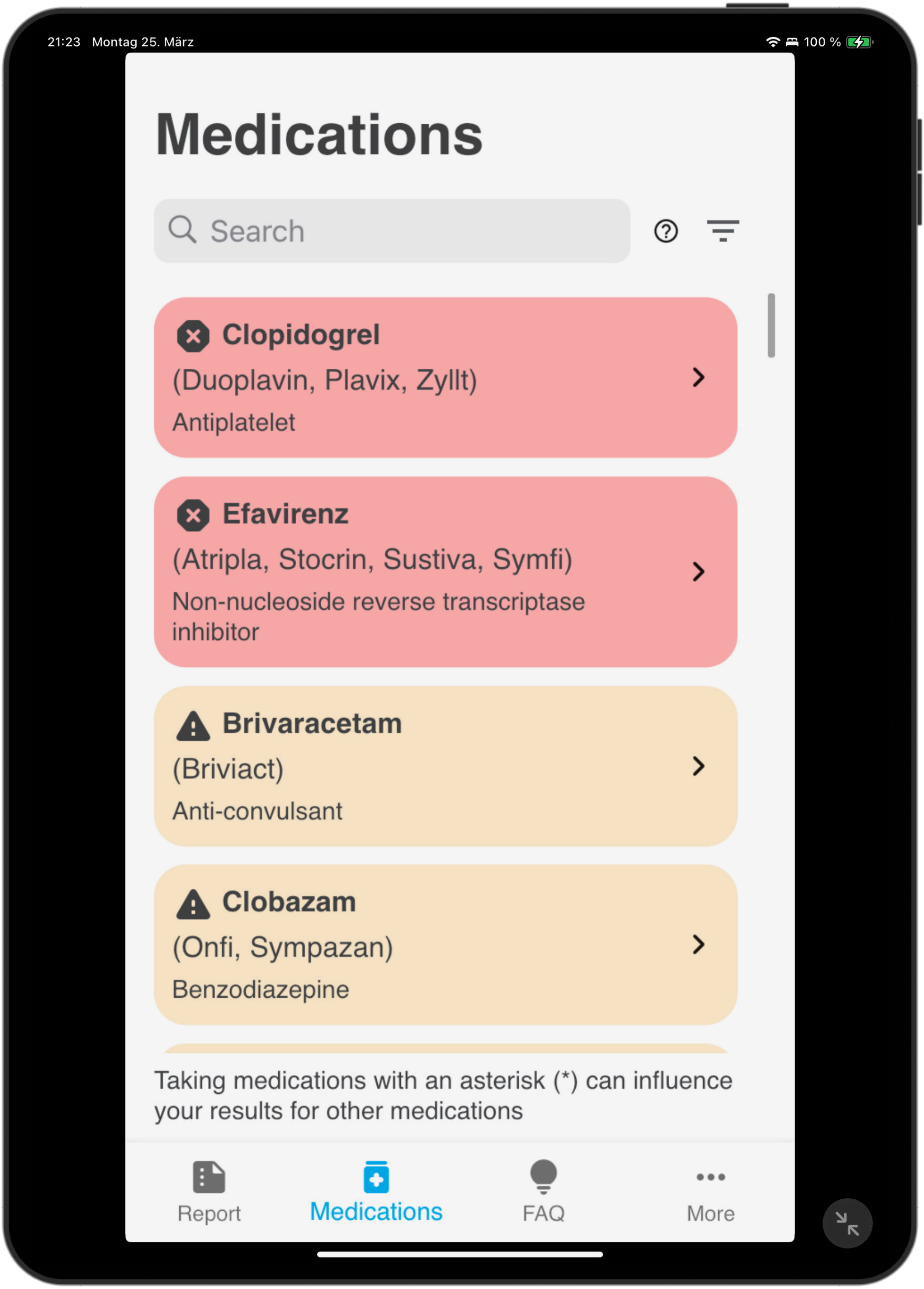
21:23 Montag 25. März
100 %
Medications
Search
Clopidogrel
(Duoplavin, Plavix, Zyllt)
Antiplatelet
Efavirenz
(Atripla, Stocrin, Sustiva, Symfi)
Non-nucleoside reverse transcriptase inhibitor
Brivaracetam
(Briviact)
Anti-convulsant
Clobazam
(Onfi, Sympazan)
Benzodiazepine
Taking medications with an asterisk (*) can influence your results for other medications
Report
Medications
FAQ
More

4.1

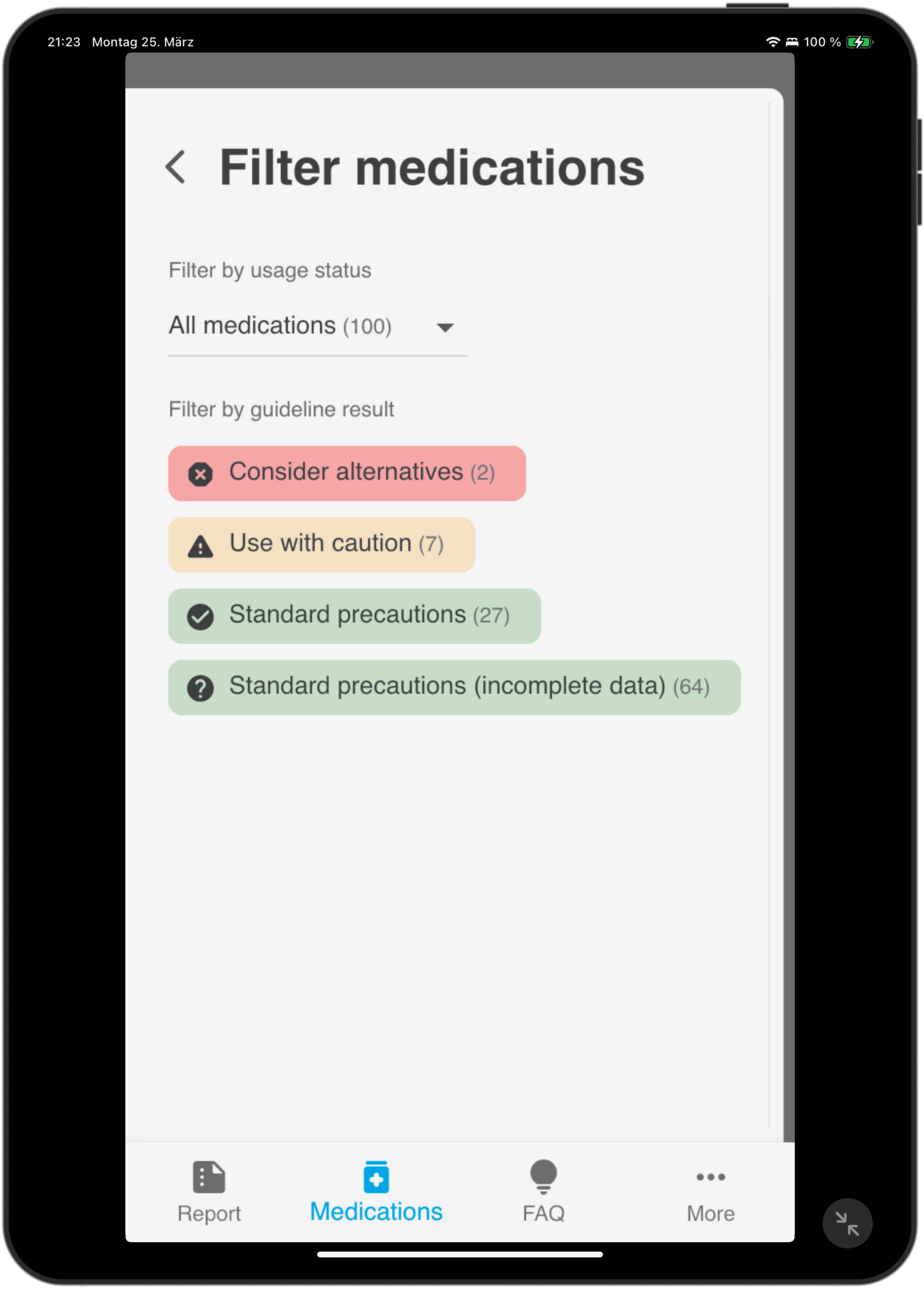
21:23 Montag 25. März
100 %
Filter medications
Filter by usage status
All medications (100)
Filter by guideline result
Consider alternatives (2)
Use with caution (7)
Standard precautions (27)
Standard precautions (incomplete data) (64)
Report
Medications
FAQ
More

4.2

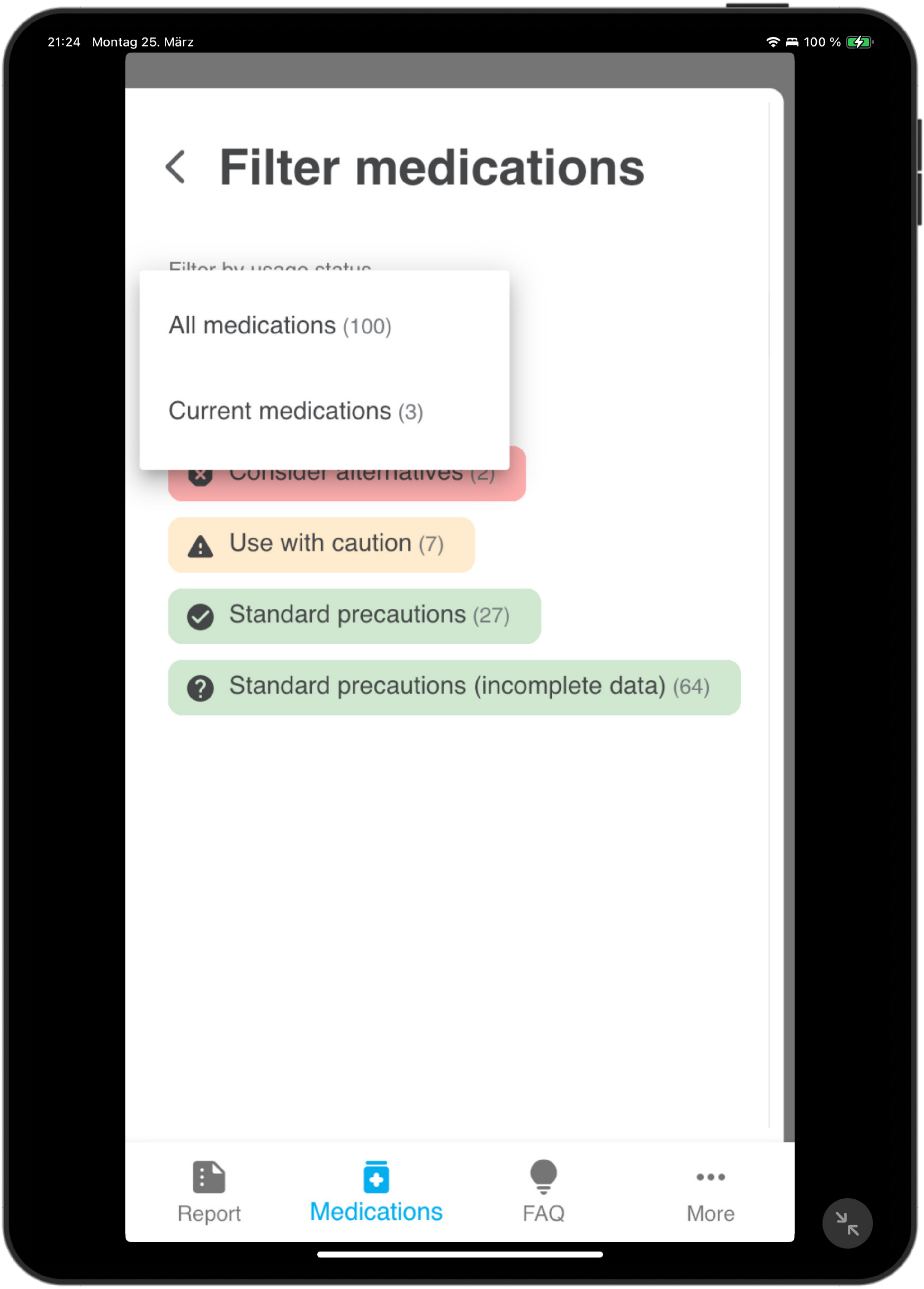
21:24 Montag 25. März
100 %
Filter medications
All medications (100)
Current medications (3)
Use with caution (7)
Standard precautions (27)
Standard precautions (incomplete data) (64)
Report
Medications
FAQ
More

4.3

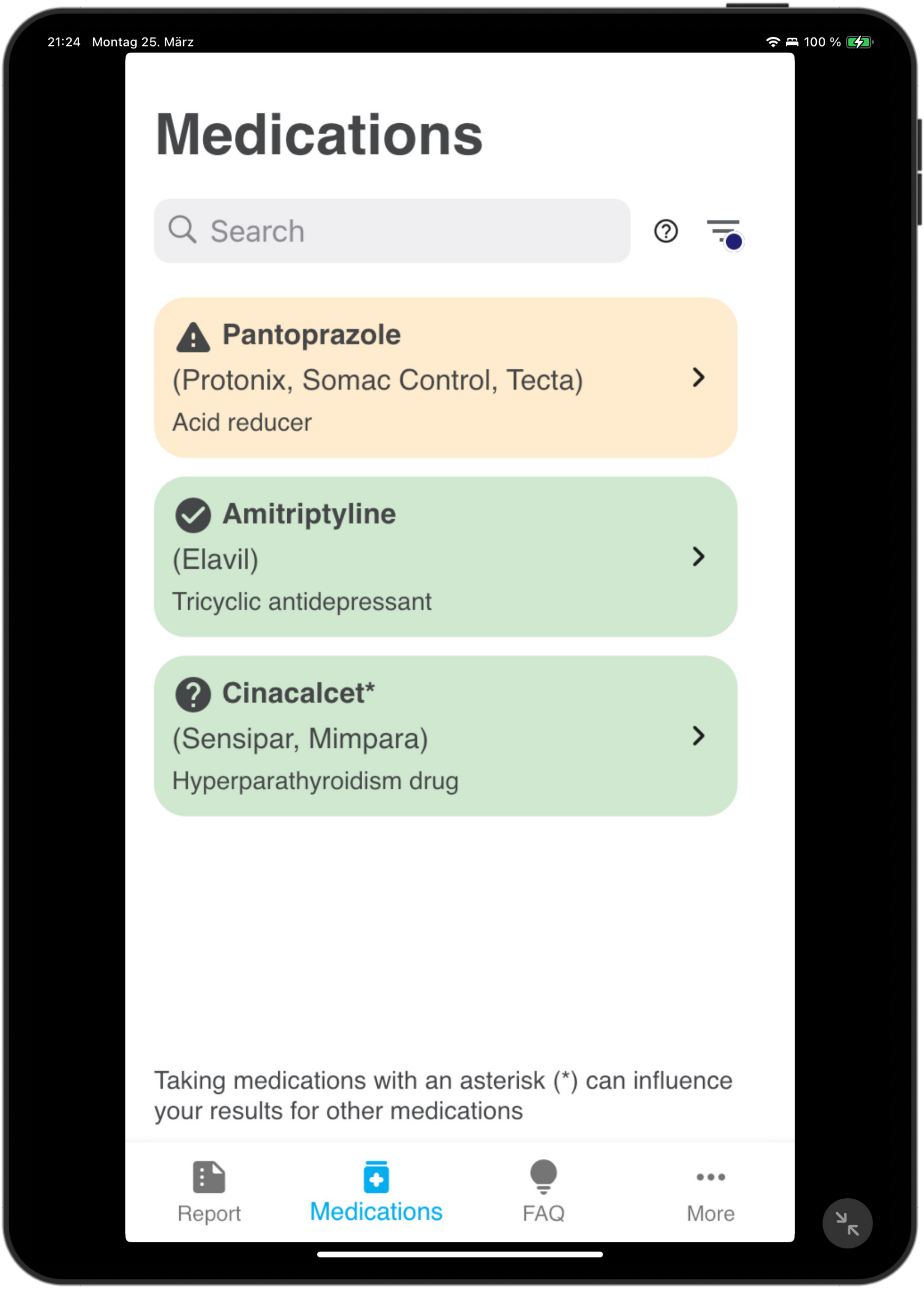
21:24 Montag 25. März
100 %
Medications
Search
Pantoprazole
(Protonix, Somac Control, Tecta)
Acid reducer
Amitriptyline
(Elavil)
Tricyclic antidepressant
Cinacalcet*
(Sensipar, Mimpara)
Hyperparathyroidism drug
Taking medications with an asterisk (*) can influence your results for other medications
Report
Medications
FAQ
More

4.4

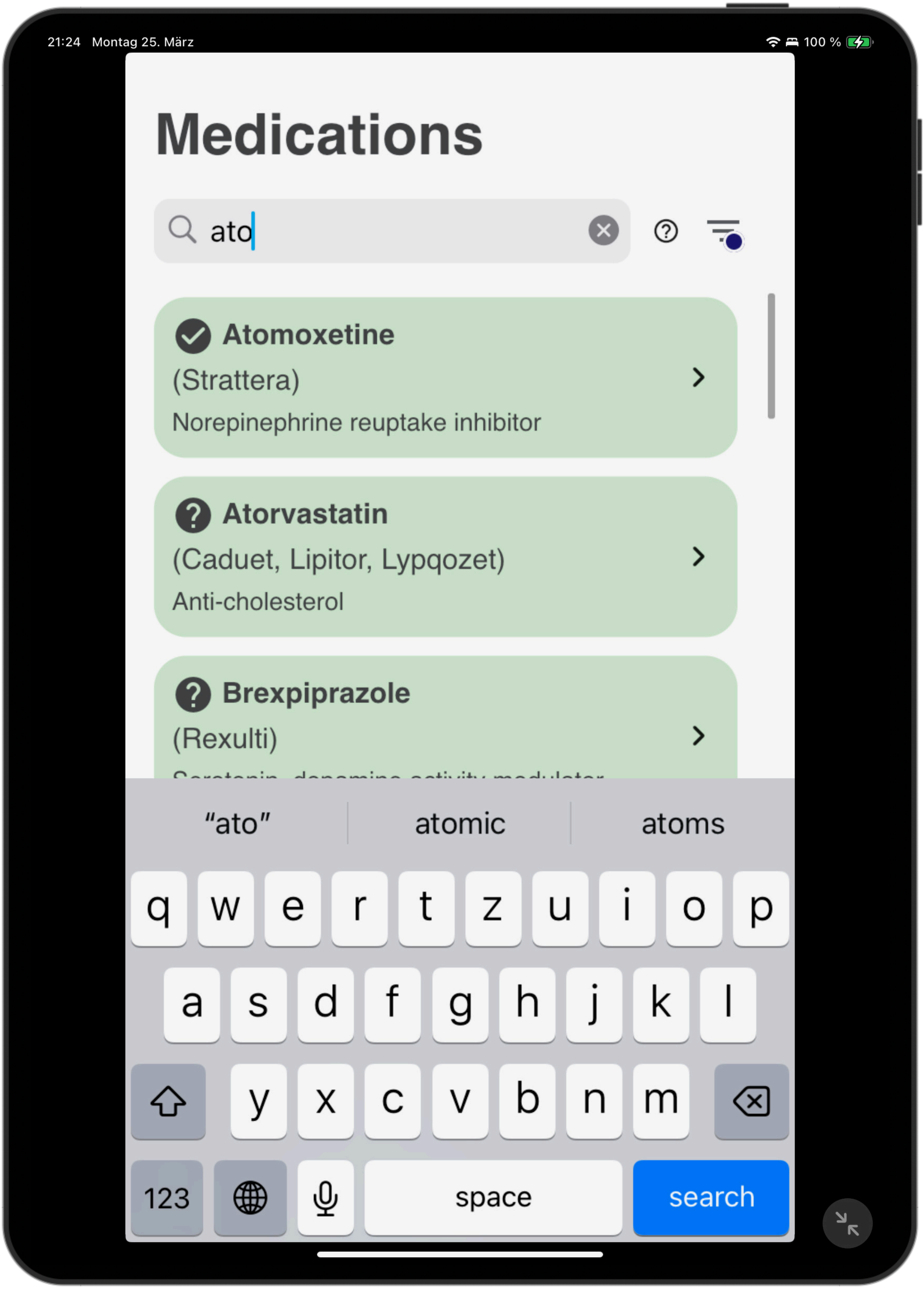
21:24 Montag 25. März
100 %
Medications
ato
Atomoxetine
(Strattera)
Norepinephrine reuptake inhibitor
Atorvastatin
(Caduet, Lipitor, Lypqozet)
Anti-cholesterol
Brexpiprazole
(Rexulti)
“ato”
atomic
atoms
q w e r t z u i o p
a s d f g h j k l
y x c v b n m
123
space
search

4.5

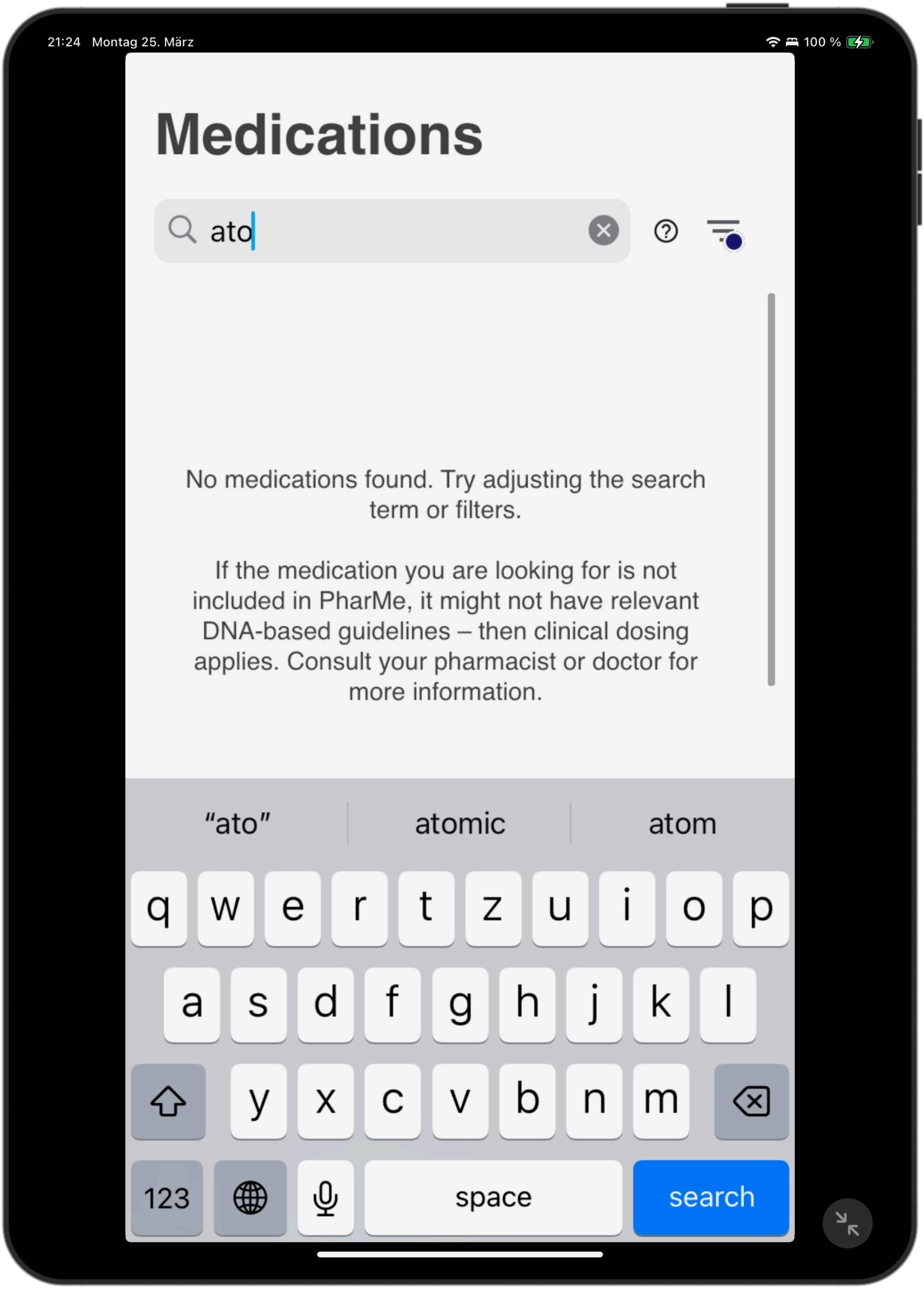
21:24 Montag 25. März
100 %
Medications
ato
No medications found. Try adjusting the search term or filters.
If the medication you are looking for is not included in PharMe, it might not have relevant DNA-based guidelines – then clinical dosing applies. Consult your pharmacist or doctor for more information.
“ato”
atomic
atom
q w e r t z u i o p
a s d f g h j k l
y x c v b n m
123
space
search

5.1

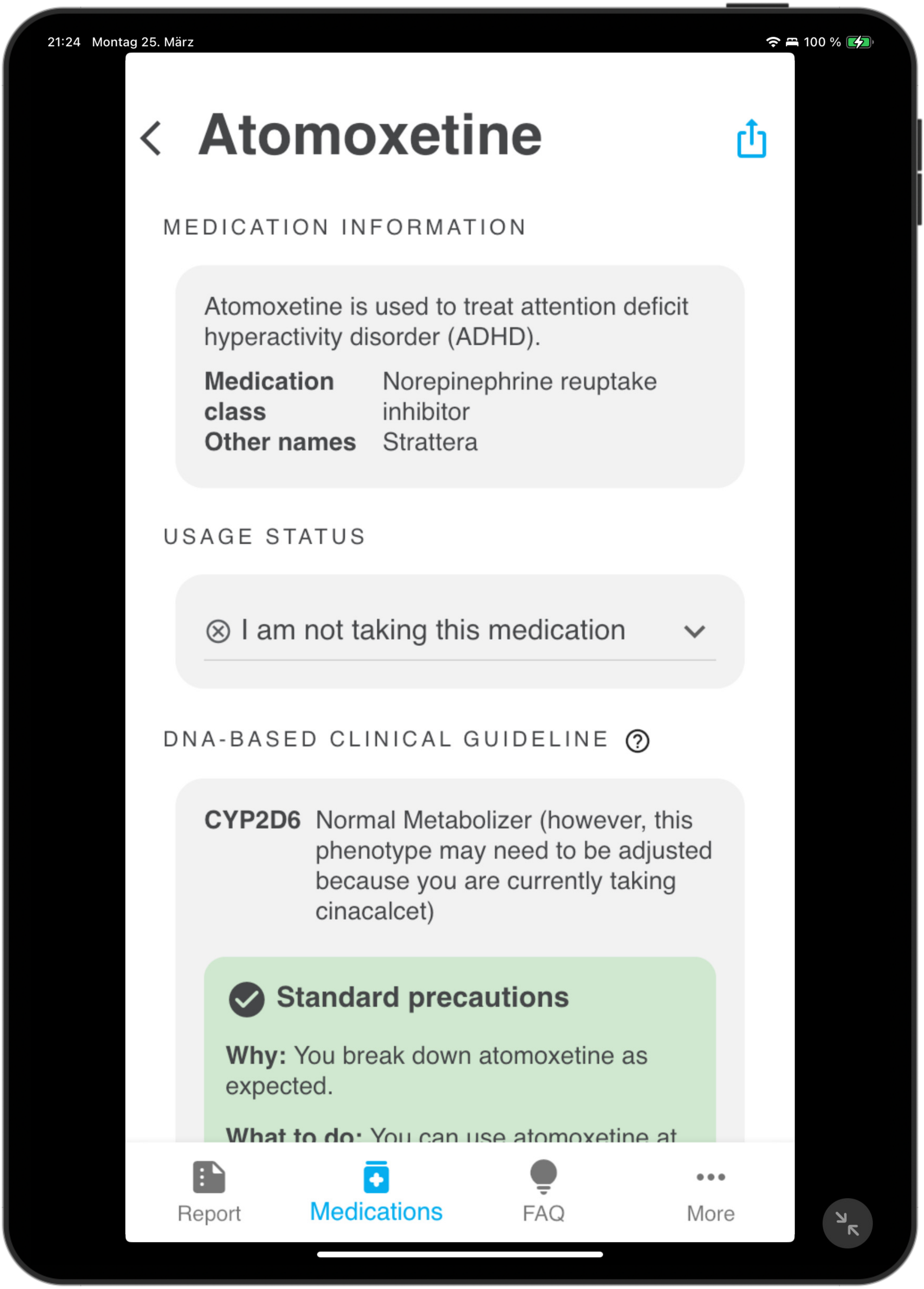
21:24 Montag 25. März
100 %
Atomoxetine
MEDICATION INFORMATION
Atomoxetine is used to treat attention deficit hyperactivity disorder (ADHD).
Medication class
Norepinephrine reuptake inhibitor
Other names
Strattera
USAGE STATUS
I am not taking this medication
DNA-BASED CLINICAL GUIDELINE
CYP2D6
Normal Metabolizer (however, this phenotype may need to be adjusted because you are currently taking cinacalcet)
Standard precautions
Why: You break down atomoxetine as expected.
Report
Medications
FAQ
More

5.2

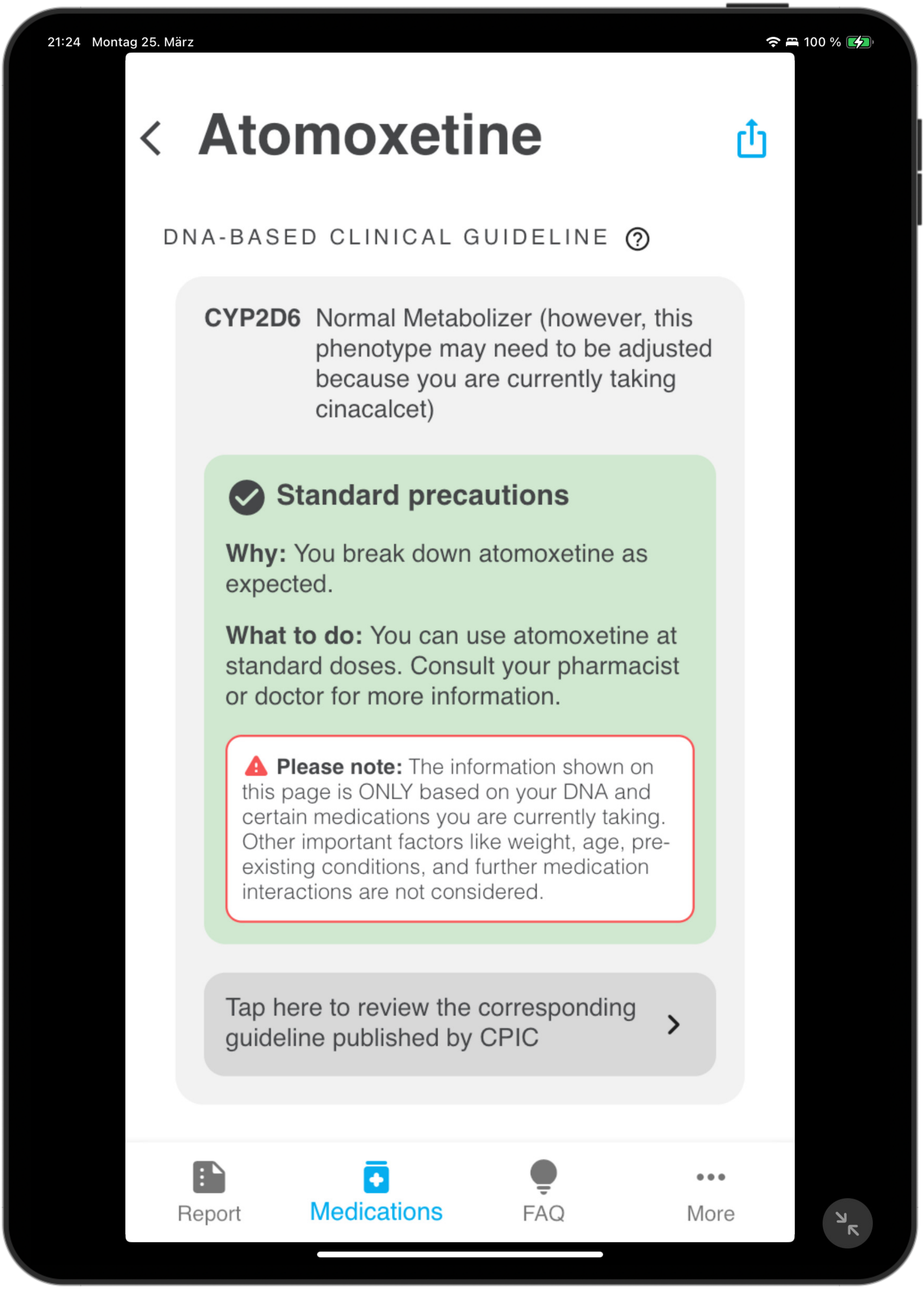
21:24 Montag 25. März
100 %
Atomoxetine
DNA-BASED CLINICAL GUIDELINE
CYP2D6 Normal Metabolizer (however, this phenotype may need to be adjusted because you are currently taking cinacalcet)
Standard precautions
Why: You break down atomoxetine as expected.
What to do: You can use atomoxetine at standard doses. Consult your pharmacist or doctor for more information.
Please note: The information shown on this page is ONLY based on your DNA and certain medications you are currently taking. Other important factors like weight, age, pre-existing conditions, and further medication interactions are not considered.
Tap here to review the corresponding guideline published by CPIC
Report
Medications
FAQ
More

6.1.1

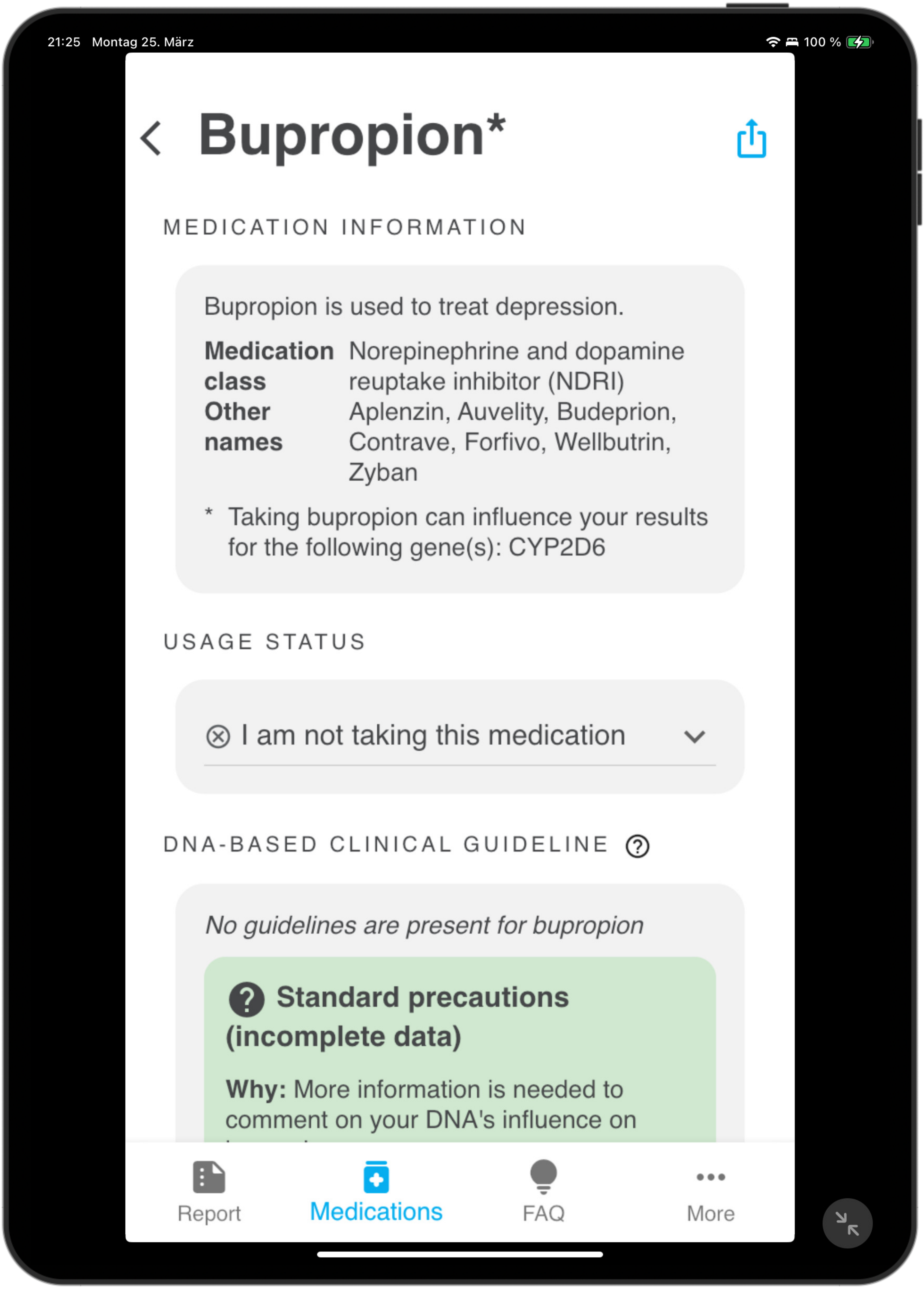
21:25 Montag 25. März
100 %
Bupropion*
MEDICATION INFORMATION
Bupropion is used to treat depression.
Medication class
Norepinephrine and dopamine reuptake inhibitor (NDRI)
Other names
Aplenzin, Auvelity, Budeprion, Contrave, Forfivo, Wellbutrin, Zyban
* Taking bupropion can influence your results for the following gene(s): CYP2D6
USAGE STATUS
I am not taking this medication
DNA-BASED CLINICAL GUIDELINE
No guidelines are present for bupropion
Standard precautions (incomplete data)
Why: More information is needed to comment on your DNA's influence on
Report
Medications
FAQ
More

6.1.2

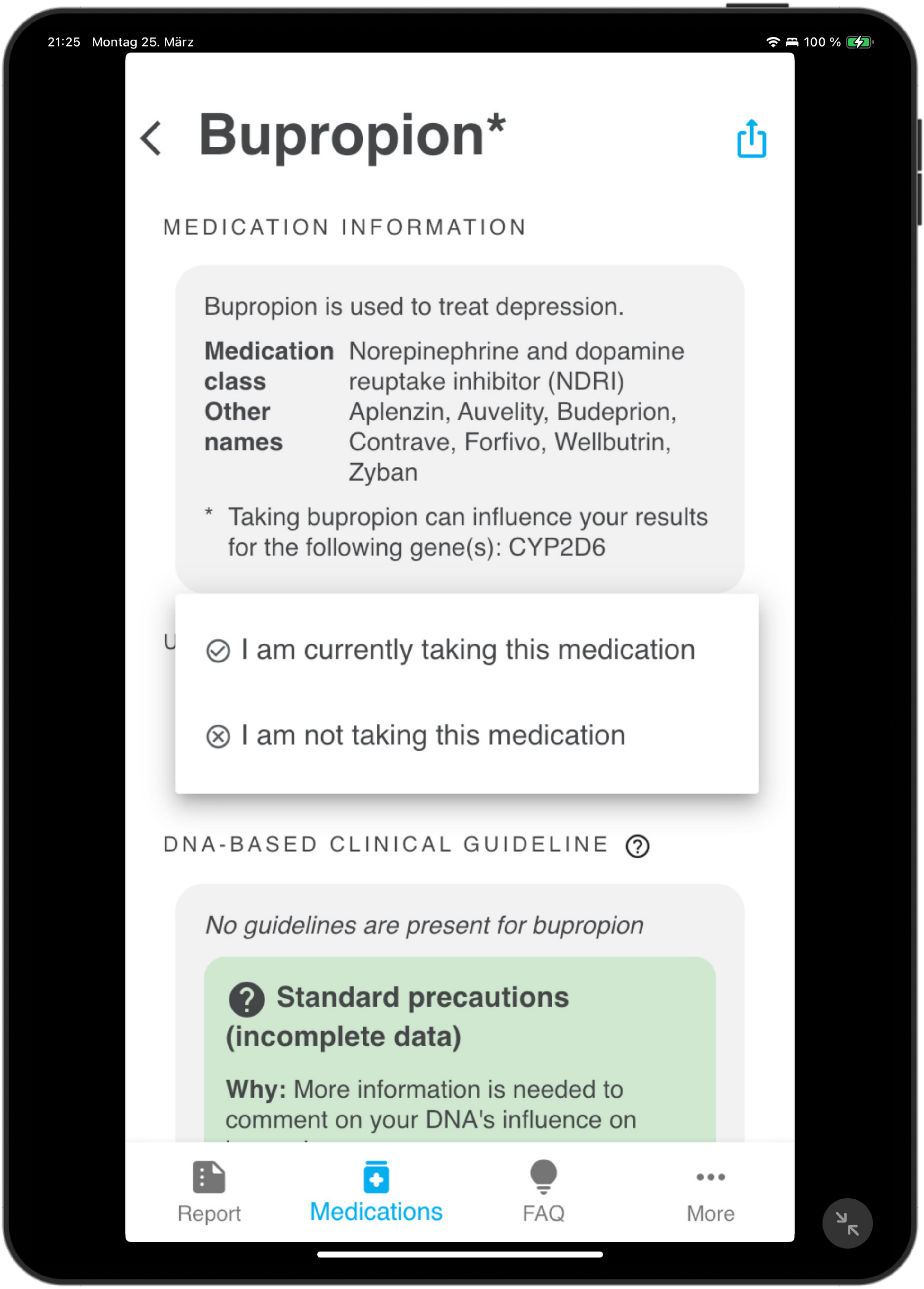
21:25 Montag 25. März
100 %
Bupropion*
MEDICATION INFORMATION
Bupropion is used to treat depression.
Medication class Norepinephrine and dopamine reuptake inhibitor (NDRI)
Other names Aplenzin, Auvelity, Budeprion, Contrave, Forfivo, Wellbutrin, Zyban
* Taking bupropion can influence your results for the following gene(s): CYP2D6
I am currently taking this medication
I am not taking this medication
DNA-BASED CLINICAL GUIDELINE
No guidelines are present for bupropion
Standard precautions (incomplete data)
Why: More information is needed to comment on your DNA's influence on
Report
Medications
FAQ
More

6.1.3

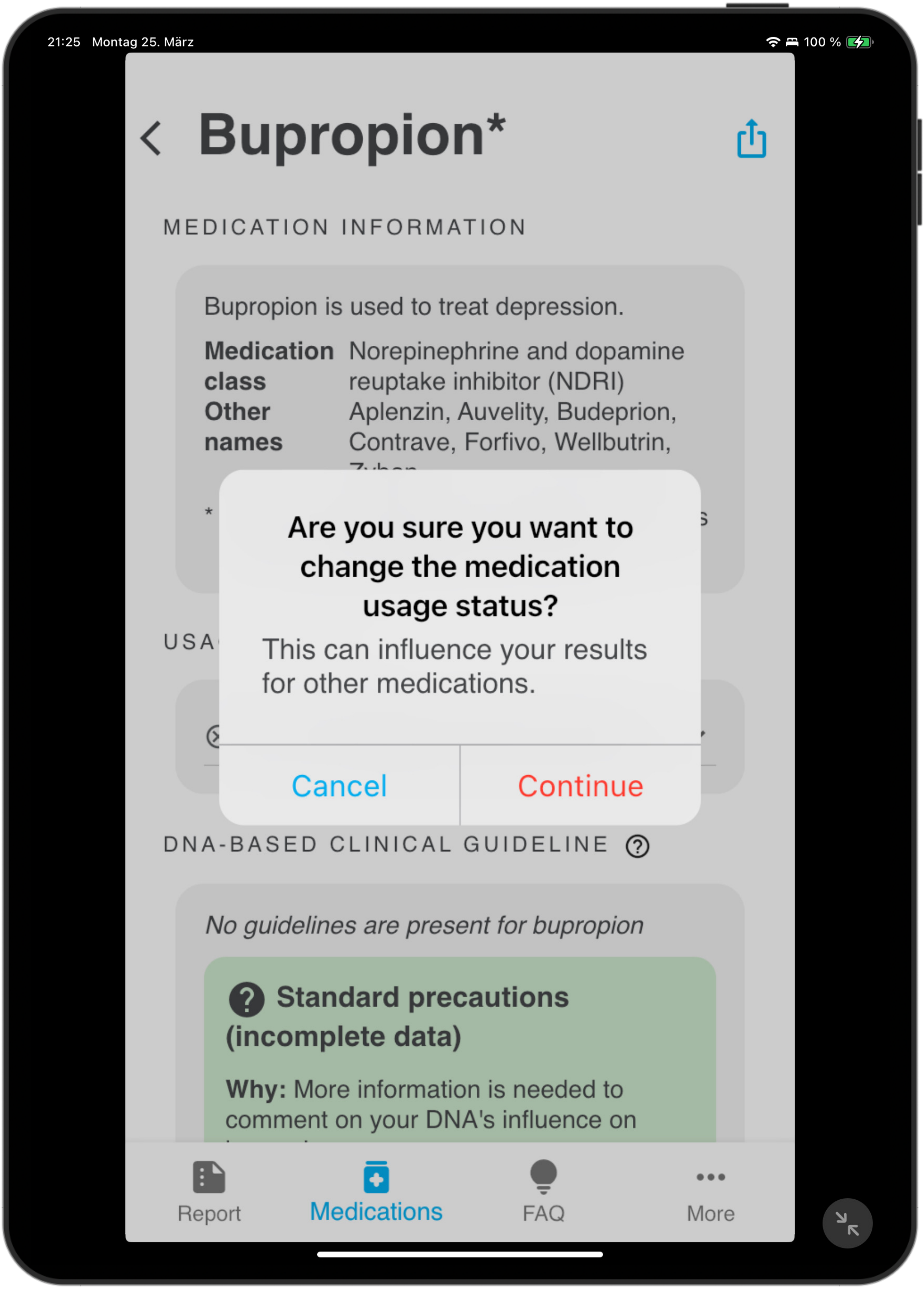

21:25 Montag 25. März
100 %
Bupropion*
MEDICATION INFORMATION
Bupropion is used to treat depression.
Medication class
Norepinephrine and dopamine reuptake inhibitor (NDRI)
Other names
Aplenzin, Auvelity, Budeprion, Contrave, Forfivo, Wellbutrin,
Are you sure you want to change the medication usage status?
This can influence your results for other medications.
Cancel
Continue
DNA-BASED CLINICAL GUIDELINE
No guidelines are present for bupropion
Standard precautions (incomplete data)
Why: More information is needed to comment on your DNA's influence on
Report
Medications
FAQ
More

6.1.4

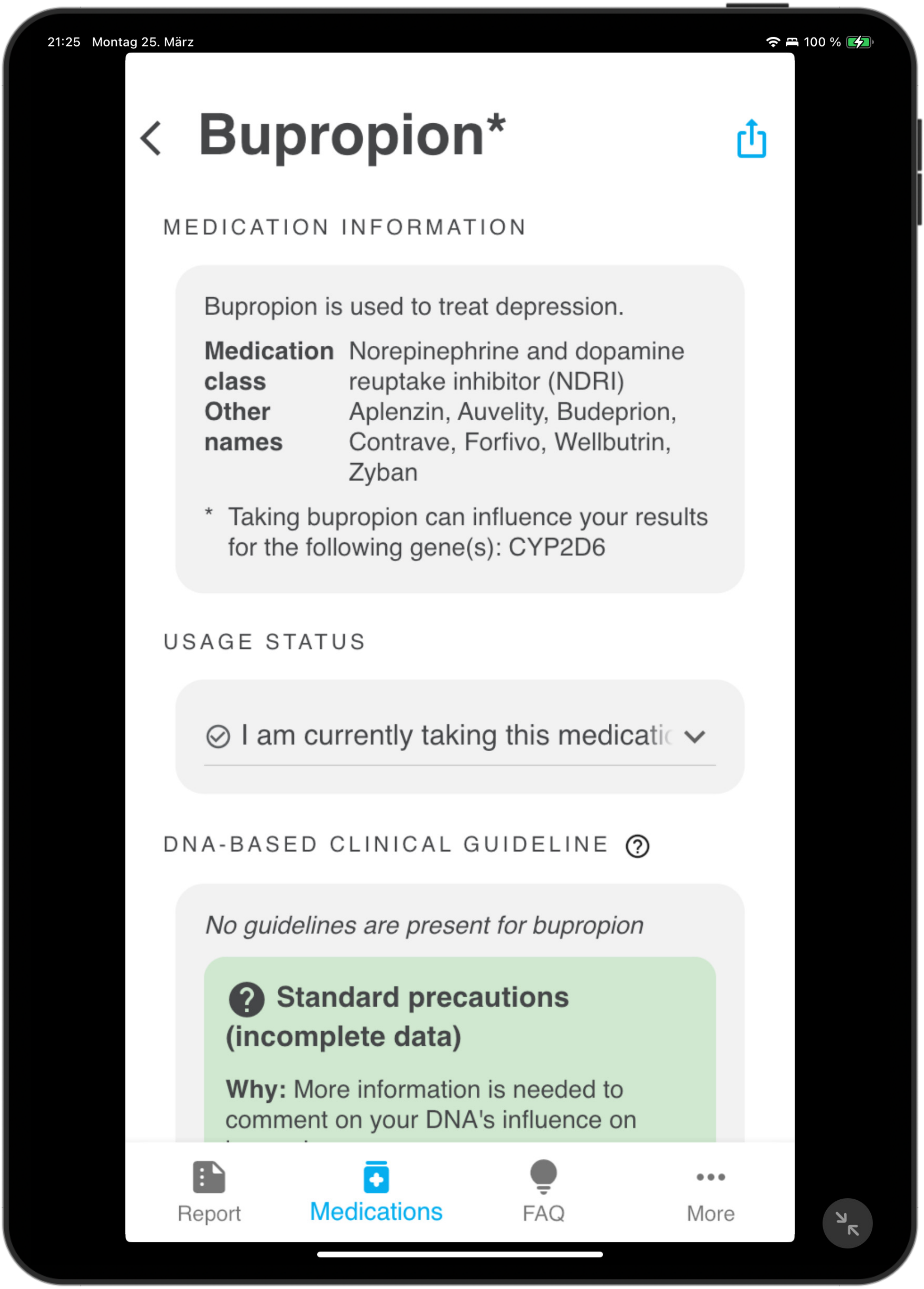
21:25 Montag 25. März
100 %
Bupropion*
MEDICATION INFORMATION
Bupropion is used to treat depression.
Medication class
Norepinephrine and dopamine reuptake inhibitor (NDRI)
Other names
Aplenzin, Auvelity, Budeprion, Contrave, Forfivo, Wellbutrin, Zyban
* Taking bupropion can influence your results for the following gene(s): CYP2D6
USAGE STATUS
I am currently taking this medicati
DNA-BASED CLINICAL GUIDELINE
No guidelines are present for bupropion
Standard precautions (incomplete data)
Why: More information is needed to comment on your DNA's influence on
Report
Medications
FAQ
More

6.2.1

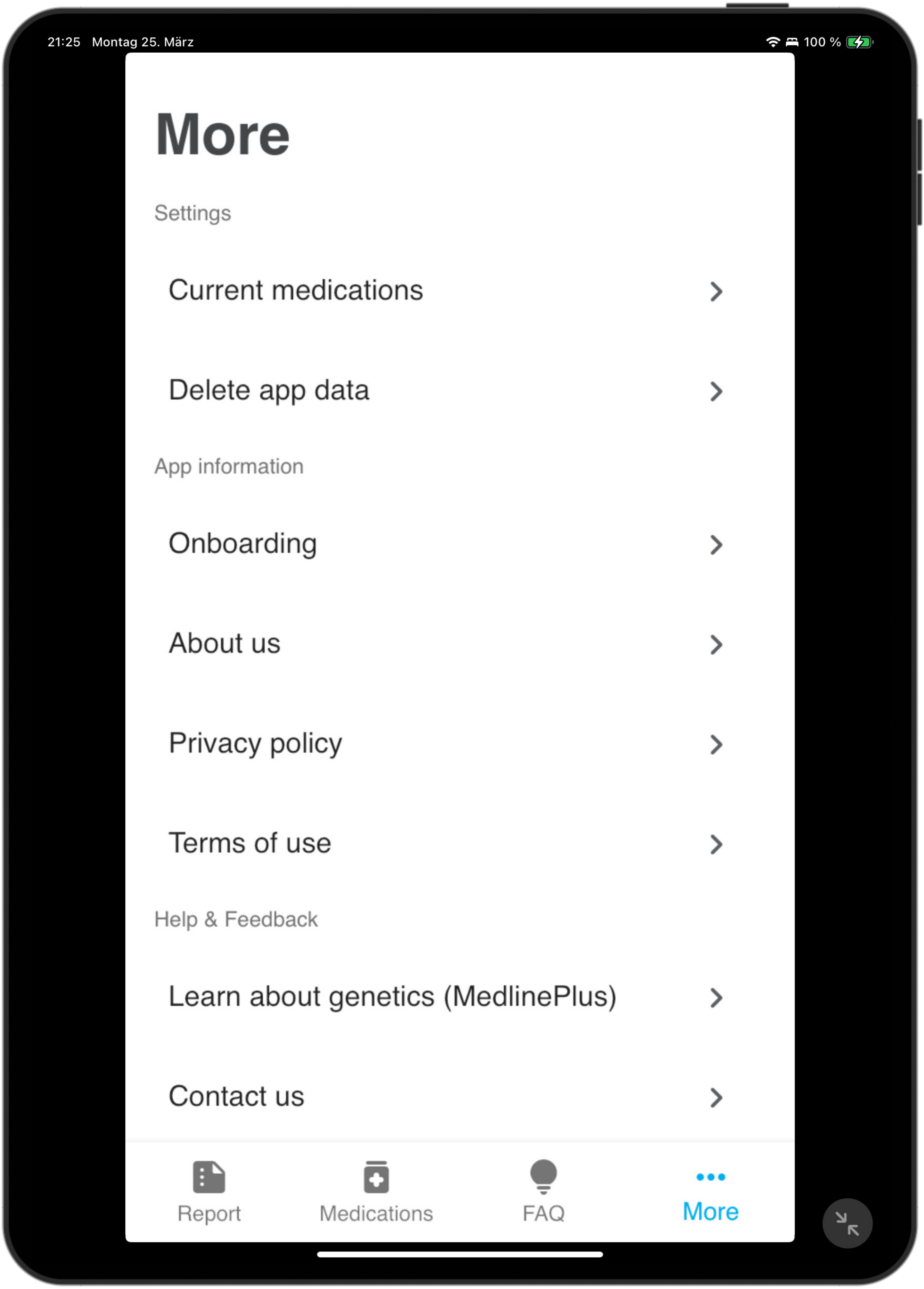
21:25 Montag 25. März
100 %
More
Settings
Current medications
Delete app data
App information
Onboarding
About us
Privacy policy
Terms of use
Help & Feedback
Learn about genetics (MedlinePlus)
Contact us
Report
Medications
FAQ
More

6.2.2

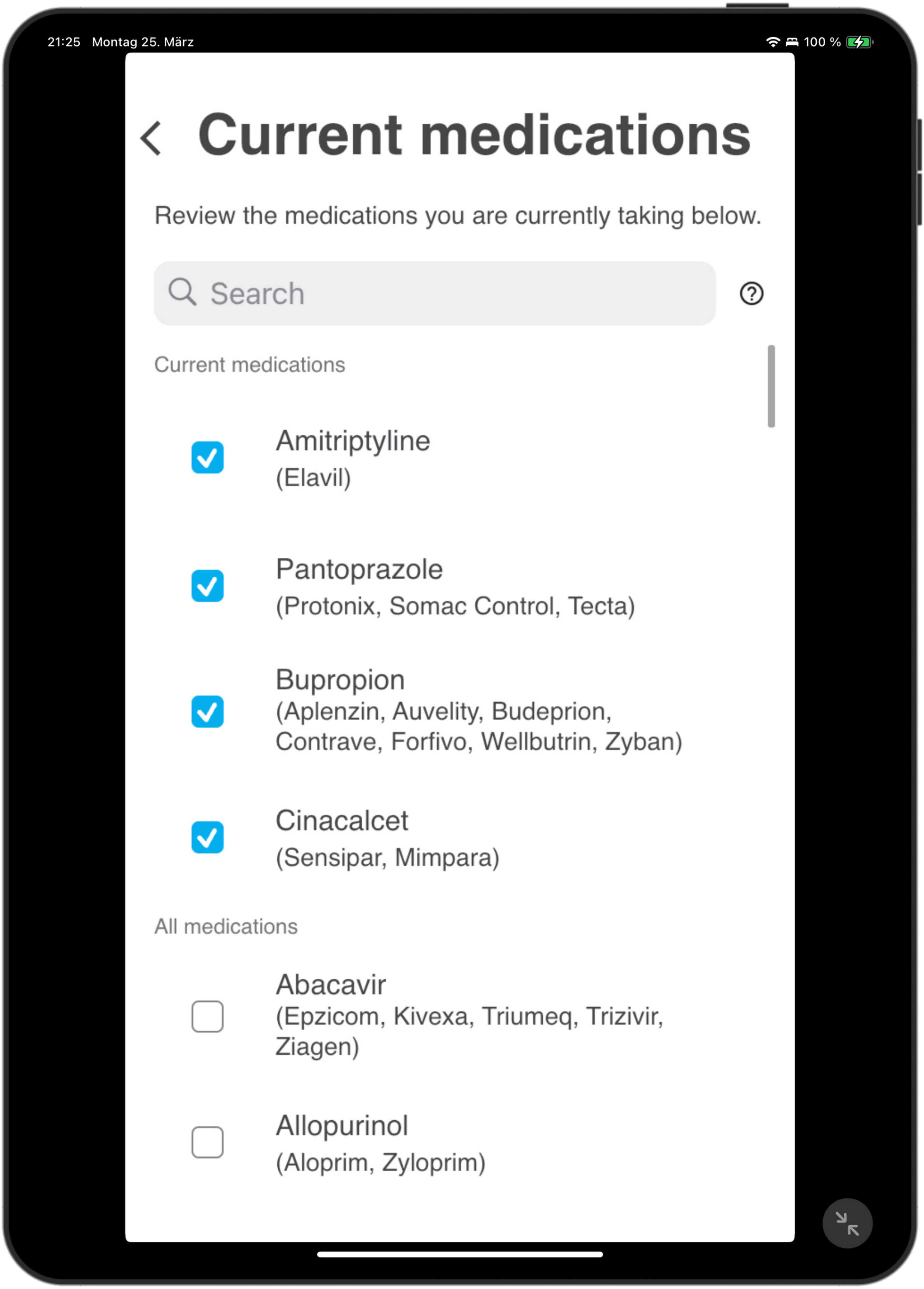
21:25 Montag 25. März
100 %
Current medications
Review the medications you are currently taking below.
Search
Current medications
Amitriptyline
(Elavil)
Pantoprazole
(Protonix, Somac Control, Tecta)
Bupropion
(Aplenzin, Auvelity, Budeprion, Contrave, Forfivo, Wellbutrin, Zyban)
Cinacalcet
(Sensipar, Mimpara)
All medications
Abacavir
(Epzicom, Kivexa, Triumeq, Trizivir, Ziagen)
Allopurinol
(Aloprim, Zyloprim)

7.1

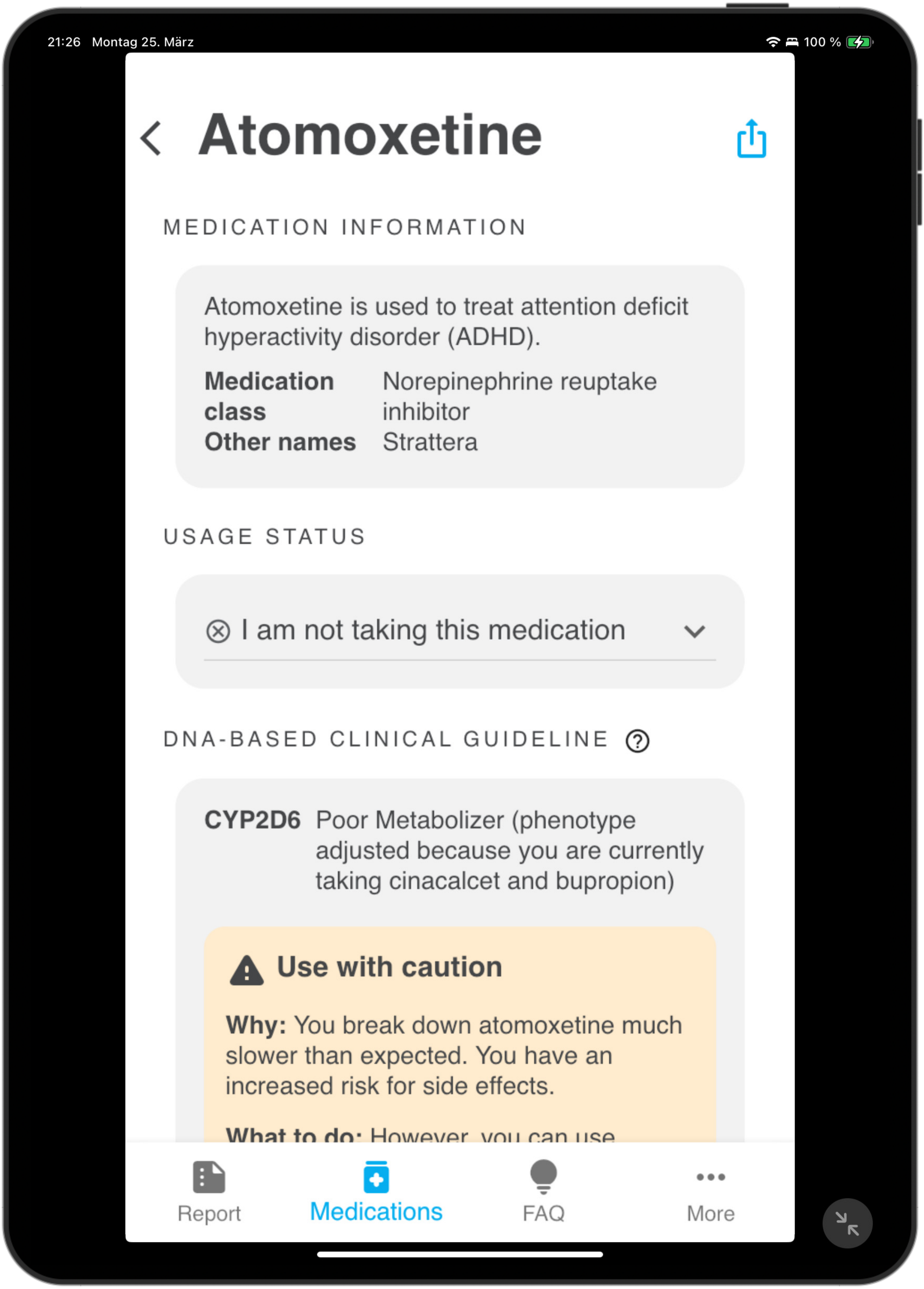
21:26 Montag 25. März
100 %
Atomoxetine
MEDICATION INFORMATION
Atomoxetine is used to treat attention deficit hyperactivity disorder (ADHD).
Medication class
Norepinephrine reuptake inhibitor
Other names
Strattera
USAGE STATUS
I am not taking this medication
DNA-BASED CLINICAL GUIDELINE
CYP2D6
Poor Metabolizer (phenotype adjusted because you are currently taking cinacalcet and bupropion)
Use with caution
Why: You break down atomoxetine much slower than expected. You have an increased risk for side effects.
Report
Medications
FAQ
More

7.1

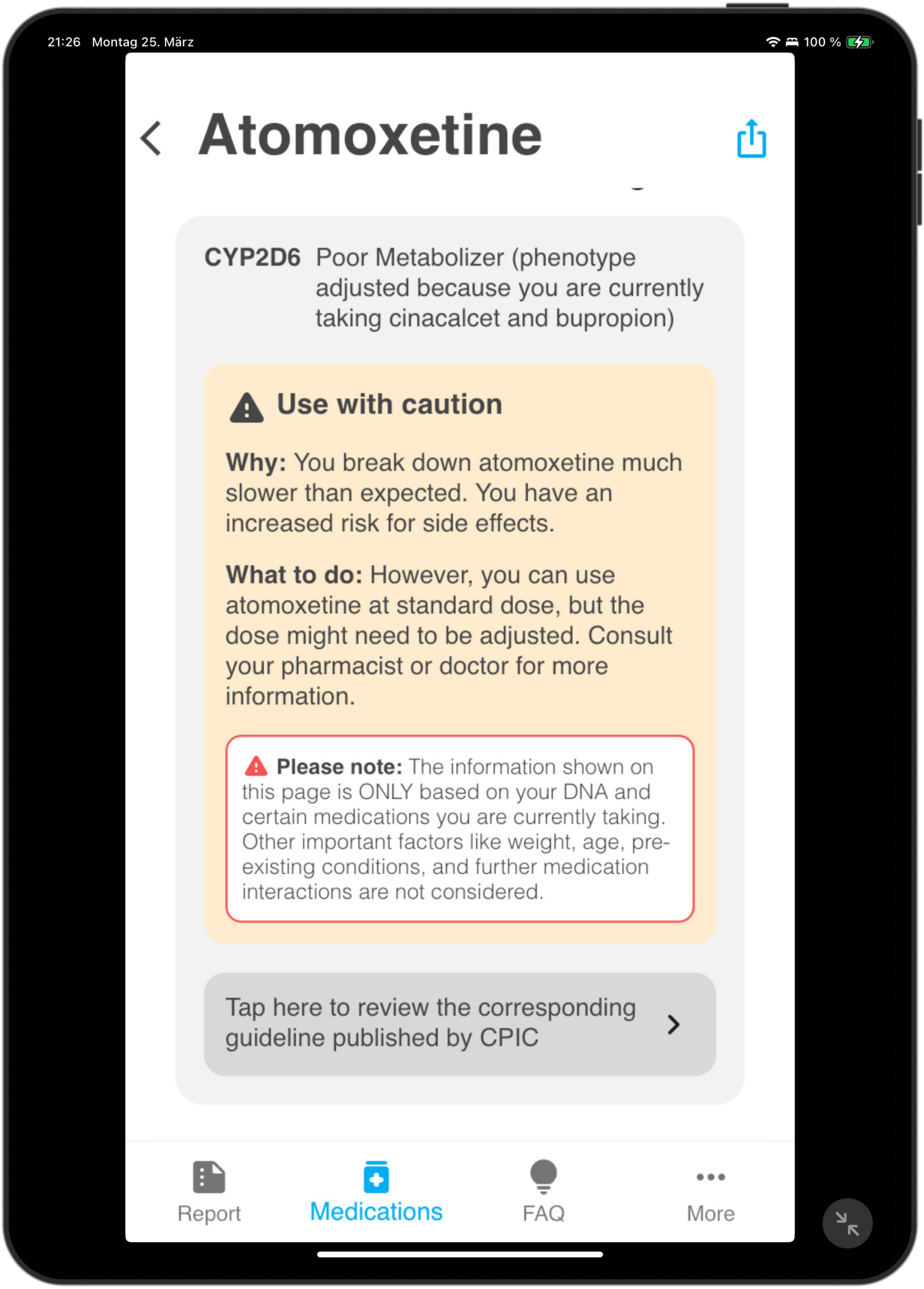
21:26 Montag 25. März
100 %
Atomoxetine
CYP2D6 Poor Metabolizer (phenotype adjusted because you are currently taking cinacalcet and bupropion)
Use with caution
Why: You break down atomoxetine much slower than expected. You have an increased risk for side effects.
What to do: However, you can use atomoxetine at standard dose, but the dose might need to be adjusted. Consult your pharmacist or doctor for more information.
Please note: The information shown on this page is ONLY based on your DNA and certain medications you are currently taking. Other important factors like weight, age, pre-existing conditions, and further medication interactions are not considered.
Tap here to review the corresponding guideline published by CPIC
Report
Medications
FAQ
More
