## Supplementary Material 3 for "Design and Implementation of PharMe: A Mobile Application to Return Pharmacogenomic Test Results to Patients"

### PharMe Pilot Testing

Thank you for testing PharMe! 🤗

The aim of the app is to provide users with pharmacogenomic (PGx) information on how their genes influence medication response, e.g., efficacy and side-effects.

In the pilot testing we want to find out which flaws PharMe still have, e.g., if you find bugs or things that are unclear. We estimate that the test will take about 15 to 20 minutes; your help would be highly appreciated!

For the test we would first like you to follow a scenario (we will explain everything step-by-step on the next pages) and answer some short questions about it. Afterwards, we welcome you to put the app through its paces and answer another short questionnaire.

Your participation is voluntary.
Your responses will be collected using Google Forms and a Google Sheet.
We do not collect identifying information.
Responses will only be visible to the study team.
We might use the collected information in publications.

We will definitely use your responses to improve the app, thank you in advance! ❤️🤝

If you have any questions or encounter anything uncertainties, please reach out to Tamara via ****. 🙏📧

* Indicates required question

1. I want to participate in the pilot testing: *

*Mark only one oval.*

- Yes
- No

#### About You

Some questions about you, so that we can get an idea of who uses PharMe in what way. 😊

(Skip questions if you prefer not so say.)

2. What is your age?

*Mark only one oval.*

- < 18 years old
- 18 - 30 years old
- 31 - 45 years old
- 46 - 60 years old
- 60+

3. What gender do you identify as?

*Mark only one oval.*

- Male
- Female
- Other

4. What is the highest degree or level of education?

*Mark only one oval.*

- Some secondary education (high school)
- Completed secondary education (graduated high school)
- Trade/technical/vocational training
- Some undergraduate education (college or university)
- Completed undergraduate education
- Some postgraduate education (masters or doctorate)
- Completed postgraduate education

#### App Testing Scenario

**Please follow the following steps to setup everything:**

1. Download the PharMe app
2. Have the login credentials you received from the study team ready
3. Open the PharMe app
4. Accept the terms and conditions
5. Log in with the credentials you received from the study team

#### Onboarding

Read through the onboarding screens until the screen you reach after logging in, until you reach the "Get started" button on the last screen.

5. Do you have any feedback for the screens? What works well? What does not work well or is unclear?

#### Initial setup

Press "Get started" to reach the medication selection. Please update the list to your following active medications:

- Pantoprazole
- Elavil
- Atomoxetine

6. Do you have any feedback for this screen? What works well? What does not work well or is unclear?

#### Tutorial

Press "Continue" to reach the tutorial; please do **not** skip. 😉

7. Do you have any feedback for the screens? What works well? What does not work well or is unclear?

#### Medications List

Everything is set up now. After finishing the tutorial, you reach the Medications screen. Please answer the questions below.

8. How can you easily only display medications you are currently taking?

9. Do you have any feedback for this screen? What works well? What does not work well or is unclear?

10. Select **atomoxetine** and review the medication information page. According to this exemplary PGx test result, if you ever needed to take the medication **atomoxetine**, could you take it at **standard dosage**?

*Mark only one oval.*

- Yes
- No

11. Do you have any feedback for this screen? What works well? What does not work well or is unclear?

#### Gene Report

Navigate to the Genes page to reach the gene report.

12. Select **CYP2D6** and review the gene information page. As best as you can, please select the phenotype for **CYP2D6**:

*Mark only one oval.*

- Ultrarapid Metabolizer
- Normal Metabolizer
- Intermediate Metabolizer
- Poor Metabolizer
- Indeterminate

13. Do you have any feedback for this screen? What works well? What does not work well or is unclear?

#### Adding a current medication

Please use PharMe to search for the medication **bupropion** now, select it, and mark it as a medication you are currently taking.

14. Do you have any feedback for this part? What works well? What does not work well or is unclear?

#### Revisiting Atomoxetine

Go back to **atomoxetine** now and review your result.

15. Can you shortly explain why the result changed?

16. Do you have any feedback for this screen? What works well? What does not work well or is unclear?

#### Testing data security features

Data security features are basically automatic logouts and hidden content if the app is not active. It would be great to see if these work everywhere – sorry for two of the tests needing waiting time, maybe grab a nice cup of tea? 🍵

17. Please pretend to switch to another app (e.g., by swiping up), so that the overview of currently opened apps shows. Does the app show only the logo and hides the app content?

*Mark only one oval.*

- ⬭ Yes
- ⬭ No

18. Please don't interact with the app for more than three minutes (e.g., use another app or leave PharMe open without doing anything); does the app prompt you to re-login?

*Mark only one oval.*

- ⬭ Yes
- ⬭ No

#### Free App Exploration

Thank you for following the scenario! Now please feel free to explore the app (e.g., more genes, medications, and the FAQ section) and answer the questions below. You can add further feedback at the end.

19. I think that I would like to use this system frequently.

*Mark only one oval.*

| | 1 | 2 | 3 | 4 | 5 | |
|---|---|---|---|---|---|---|
| Stro | ⬭ | ⬭ | ⬭ | ⬭ | ⬭ | Strongly Agree |

20. I found the system unnecessarily complex.

*Mark only one oval.*

| | 1 | 2 | 3 | 4 | 5 | |
|---|---|---|---|---|---|---|
| Stro | | | | | | Strongly Agree |

21. I thought the system was easy to use.

*Mark only one oval.*

| | 1 | 2 | 3 | 4 | 5 | |
|---|---|---|---|---|---|---|
| Stro | | | | | | Strongly Agree |

22. I think that I would need the support of a technical person to be able to use this system.

*Mark only one oval.*

| | 1 | 2 | 3 | 4 | 5 | |
|---|---|---|---|---|---|---|
| Stro | | | | | | Strongly Agree |

23. I found the various functions in this system were well integrated.

*Mark only one oval.*

| | 1 | 2 | 3 | 4 | 5 | |
|---|---|---|---|---|---|---|
| Stro | | | | | | Strongly Agree |

24. I thought there was too much inconsistency in this system.

*Mark only one oval.*

| | 1 | 2 | 3 | 4 | 5 | |
|---|---|---|---|---|---|---|
| Stro | ◯ | ◯ | ◯ | ◯ | ◯ | Strongly Agree |

25. I would imagine that most people would learn to use this system very quickly.

*Mark only one oval.*

| | 1 | 2 | 3 | 4 | 5 | |
|---|---|---|---|---|---|---|
| Stro | ◯ | ◯ | ◯ | ◯ | ◯ | Strongly Agree |

26. I found the system very cumbersome to use.

*Mark only one oval.*

| | 1 | 2 | 3 | 4 | 5 | |
|---|---|---|---|---|---|---|
| Stro | ◯ | ◯ | ◯ | ◯ | ◯ | Strongly Agree |

27. I felt very confident using the system.

*Mark only one oval.*

| | 1 | 2 | 3 | 4 | 5 | |
|---|---|---|---|---|---|---|
| Stro | ◯ | ◯ | ◯ | ◯ | ◯ | Strongly Agree |

28. I needed to learn a lot of things before I could get going with this system.

*Mark only one oval.*

| | 1 | 2 | 3 | 4 | 5 | |
|---|---|---|---|---|---|---|
| Stro | ◯ | ◯ | ◯ | ◯ | ◯ | Strongly Agree |

29. If you would rate PharMe in the app store, how many stars would you give? (Please be honest, no need to spare my feelings 😉)

*Mark only one oval.*

| | 1 | 2 | 3 | 4 | 5 | |
|---|---|---|---|---|---|---|
| ⭐ | ◯ | ◯ | ◯ | ◯ | ◯ | ⭐⭐⭐⭐⭐ |

30. Please explain your star rating shortly.

_______________________________________________

_______________________________________________

_______________________________________________

_______________________________________________

_______________________________________________

31. Do you have any more feedback for us? What works well? What does not work well or is unclear?

_______________________________________________

_______________________________________________

_______________________________________________

_______________________________________________

_______________________________________________

This content is neither created nor endorsed by Google.

Google Forms
