## Supplementary Material 5 for "Design and Implementation of PharMe: A Mobile Application to Return Pharmacogenomic Test Results to Patients"

### *Included Medications and Genes*

| ***Medication*** | ***Actionable Genes*** | | | | ***Clinical Specialty*** |
|---|---|---|---|---|---|
| | ***Gene 1*** | ***Gene 2*** | ***Gene 3*** | ***Gene 4*** | |
| *Abrocitinib* | *CYP2C19* | | | | *Dermatology* |
| *Amphetamine* | *CYP2D6* | | | | *Behavioral Health* |
| *Aripiprazole* | *CYP2D6* | | | | *Behavioral Health* |
| *Belinostat* | *UGT1A1* | | | | *Hematology / Oncology* |
| *Belzutifan* | *CYP2C19* | | | | *Hematology / Oncology* |
| *Brexpiprazole* | *CYP2D6* | | | | *Behavioral Health* |
| *Brivaracetam* | *CYP2C19* | | | | *Neurology* |
| *Cevimeline* | *CYP2D6* | | | | *Miscellaneous* |
| *Clobazam* | *CYP2C19* | | | | *Neurology* |
| *Clozapine* | *CYP2D6* | | | | *Behavioral Health* |
| *Deutetrabenazine* | *CYP2D6* | | | | *Neurology* |
| *Dronabinol* | *CYP2C9* | | | | *Gastroenterology* |
| *Eliglustat* | *CYP2D6* | | | | *Miscellaneous* |
| *Erdafitinib* | *CYP2C9* | | | | *Hematology / Oncology* |
| *Flibanserin* | *CYP2C19* | | | | *Reproductive and Sexual Health* |
| *Gefitinib* | *CYP2D6* | | | | *Hematology / Oncology* |
| *Iloperidone* | *CYP2D6* | | | | *Behavioral Health* |
| *Irinotecan* | *UGT1A1* | | | | *Hematology / Oncology* |
| *Lofexidine* | *CYP2D6* | | | | *Behavioral Health* |
| *Mavacamten* | *CYP2C19* | | | | *Cardiology* |
| *Meclizine* | *CYP2D6* | | | | *Gastroenterology* |

| | | | | | |
|---|---|---|---|---|---|
| *Metoclopramide* | *CYP2D6* | | | | *Gastroenterology* |
| *Nateglinide* | *CYP2C9* | | | | *Anti-Diabetic* |
| *Nilotinib* | *UGT1A1* | | | | *Hematology / Oncology* |
| *Oliceridine* | *CYP2D6* | | | | *Pain Management* |
| *Pazopanib* | *UGT1A1* | *HLA-B*57:01* | | | *Hematology / Oncology* |
| *Perphenazine* | *CYP2D6* | | | | *Behavioral Health* |
| *Pimozide* | *CYP2D6* | | | | *Neurology* |
| *Pitolisant* | *CYP2D6* | | | | *Neurology* |
| *Propafenone* | *CYP2D6* | | | | *Cardiology* |
| *Sacituzumab Govitecan-hziy* | *UGT1A1* | | | | *Hematology / Oncology* |
| *Siponimod* | *CYP2C9* | | | | *Neurology* |
| *Tetrabenazine* | *CYP2D6* | | | | *Neurology* |
| *Thioridazine* | *CYP2D6* | | | | *Behavioral Health* |
| *Tolterodine* | *CYP2D6* | | | | *Urology* |
| *Valbenazine* | *CYP2D6* | | | | *Neurology* |
| *Atomoxetine* | *CYP2D6* | | | | *Behavioral Health* |
| *Acebutolol** | *CYP2D6* | | | | *Cardiology* |
| *Betaxolol** | *CYP2D6* | | | | *Cardiology* |
| *Bisoprolol** | *CYP2D6* | | | | *Cardiology* |
| *Carvedilol** | *CYP2D6* | | | | *Cardiology* |
| *Metoprolol* | *CYP2D6* | | | | *Cardiology* |
| *Nebivolol** | *CYP2D6* | | | | *Cardiology* |
| *Propranolol** | *CYP2D6* | | | | *Cardiology* |
| *Efavirenz* | *CYP2B6* | | | | *Infectious Diseases* |
| *Aminosalicylic acid** | *G6PD* | | | | *Infectious Diseases* |

| | | | | | |
|---|---|---|---|---|---|
| *Aspirin** | *G6PD* | | | | *Pain Management / Cardiology* |
| *Chloramphenicol** | *G6PD* | | | | *Infectious Diseases* |
| *Chloroquine** | *G6PD* | | | | *Infectious Diseases* |
| *Chlorpropamide** | *G6PD* | | | | *Anti-Diabetic* |
| *Ciprofloxacin** | *G6PD* | | | | *Infectious Diseases* |
| *Dabrafenib** | *G6PD* | | | | *Hematology / Oncology* |
| *Dapsone* | *G6PD* | | | | *Infectious Diseases* |
| *Dimercaprol** | *G6PD* | | | | *Antidote* |
| *Doxorubicin** | *G6PD* | | | | *Hematology / Oncology* |
| *Furazolidone** | *G6PD* | | | | *Infectious Diseases* |
| *Gliclazide** | *G6PD* | | | | *Anti-Diabetic* |
| *Glimepiride** | *G6PD* | | | | *Anti-Diabetic* |
| *Glipizide** | *G6PD* | | | | *Anti-Diabetic* |
| *Glyburide** | *G6PD* | | | | *Anti-Diabetic* |
| *Hydroxychloroquine** | *G6PD* | | | | *Infectious Diseases* |
| *Mafenide** | *G6PD* | | | | *Infectious Diseases* |
| *Mepacrine** | *G6PD* | | | | *Infectious Diseases* |
| *Mesalazine** | *G6PD* | | | | *Gastroenterology* |
| *Methylene blue* | *G6PD* | | | | *Antidote* |
| *Moxifloxacin** | *G6PD* | | | | *Infectious Diseases* |
| *Nalidixic acid** | *G6PD* | | | | *Infectious Diseases* |
| *Nicorandil** | *G6PD* | | | | *Cardiology* |
| *Nitrofural** | *G6PD* | | | | *Infectious Diseases* |
| *Nitrofurantoin* | *G6PD* | | | | *Infectious Diseases* |
| *Norfloxacin** | *G6PD* | | | | *Infectious Diseases* |

| | | | | | |
|---|---|---|---|---|---|
| *Ofloxacin** | *G6PD* | | | | *Infectious Diseases* |
| *Pegloticase* | *G6PD* | | | | *Rheumatology* |
| *Phenazopyridine** | *G6PD* | | | | *Infectious Diseases* |
| *Primaquine* | *G6PD* | | | | *Infectious Diseases* |
| *Probenecid** | *G6PD* | | | | *Rheumatology* |
| *Quinine** | *G6PD* | | | | *Infectious Diseases* |
| *Rasburicase* | *G6PD* | | | | *Hematology / Oncology* |
| *Sodium nitrite** | *G6PD* | | | | *Antidote* |
| *Sulfacetamide** | *G6PD* | | | | *Infectious Diseases* |
| *Sulfadiazine** | *G6PD* | | | | *Infectious Diseases* |
| *Sulfadimidine** | *G6PD* | | | | *Infectious Diseases* |
| *Sulfamethoxazole / trimethoprim** | *G6PD* | | | | *Infectious Diseases* |
| *Sulfanilamide** | *G6PD* | | | | *Infectious Diseases* |
| *Sulfasalazine** | *G6PD* | | | | *Gastroenterology / Rheumatology* |
| *Sulfisoxazole** | *G6PD* | | | | *Infectious Diseases* |
| *Tafenoquine* | *G6PD* | | | | *Infectious Diseases* |
| *Tolazamide** | *G6PD* | | | | *Anti-Diabetic* |
| *Tolbutamide** | *G6PD* | | | | *Anti-Diabetic* |
| *Toluidine blue* | *G6PD* | | | | *Miscellaneous* |
| *Trametinib** | *G6PD* | | | | *Hematology / Oncology* |
| *Vitamin C** | *G6PD* | | | | *Miscellaneous* |
| *Vitamin K** | *G6PD* | | | | *Miscellaneous* |
| *Aceclofenac** | *CYP2C9* | | | | *Pain Management* |
| *Celecoxib* | *CYP2C9* | | | | *Pain Management* |
| *Diclofenac** | *CYP2C9* | | | | *Pain Management* |

| | | | | | |
|---|---|---|---|---|---|
| *Flurbiprofen* | *CYP2C9* | | | | *Pain Management* |
| *Ibuprofen* | *CYP2C9* | | | | *Pain Management* |
| *Indomethacin** | *CYP2C9* | | | | *Pain Management* |
| *Lornoxicam* | *CYP2C9* | | | | *Pain Management* |
| *Meloxicam* | *CYP2C9* | | | | *Pain Management* |
| *Nabumetone** | *CYP2C9* | | | | *Pain Management* |
| *Naproxen** | *CYP2C9* | | | | *Pain Management* |
| *Piroxicam* | *CYP2C9* | | | | *Pain Management* |
| *Tenoxicam* | *CYP2C9* | | | | *Pain Management* |
| *Dexlansoprazole* | *CYP2C19* | | | | *Gastroenterology* |
| *Esomeprazole** | *CYP2C19* | | | | *Gastroenterology* |
| *Lansoprazole* | *CYP2C19* | | | | *Gastroenterology* |
| *Omeprazole* | *CYP2C19* | | | | *Gastroenterology* |
| *Pantoprazole* | *CYP2C19* | | | | *Gastroenterology* |
| *Rabeprazole** | *CYP2C19* | | | | *Gastroenterology* |
| *Citalopram* | *CYP2C19* | | | | *Behavioral Health* |
| *Duloxetine** | *CYP2C19* | *CYP2D6* | | | *Behavioral Health* |
| *Escitalopram* | *CYP2C19* | | | | *Behavioral Health* |
| *Fluoxetine** | *CYP2C19* | *CYP2D6* | | | *Behavioral Health* |
| *Fluvoxamine* | *CYP2D6* | | | | *Behavioral Health* |
| *Paroxetine* | *CYP2D6* | | | | *Behavioral Health* |
| *Sertraline* | *CYP2C19* | *CYP2B6* | | | *Behavioral Health* |
| *Venlafaxine* | *CYP2D6* | | | | *Behavioral Health* |
| *Vortioxetine* | *CYP2D6* | | | | *Behavioral Health* |
| *Atorvastatin* | *SLCO1B1* | | | | *Cardiology* |

| | | | | | |
|---|---|---|---|---|---|
| *Fluvastatin* | *SLCO1B1* | *CYP2C9* | | | *Cardiology* |
| *Lovastatin* | *SLCO1B1* | | | | *Cardiology* |
| *Pitavastatin* | *SLCO1B1* | | | | *Cardiology* |
| *Pravastatin* | *SLCO1B1* | | | | *Cardiology* |
| *Rosuvastatin* | *SLCO1B1* | *ABCG2* | | | *Cardiology* |
| *Simvastatin* | *SLCO1B1* | | | | *Cardiology* |
| *Tamoxifen* | *CYP2D6* | | | | *Hematology / Oncology* |
| *Abacavir* | *HLA-B*57:01* | | | | *Infectious Diseases* |
| *Allopurinol* | *HLA-B*58:01* | | | | *Rheumatology* |
| *Atazanavir* | *UGT1A1* | | | | *Infectious Diseases* |
| *Carbamazepine* | *HLA-B*15:02* | *HLA-A*31:01* | | | *Neurology* |
| *Oxcarbazepine* | *HLA-B*15:02* | | | | *Neurology* |
| *Clopidogrel* | *CYP2C19* | | | | *Cardiology* |
| *Codeine* | *CYP2D6* | | | | *Pain Management* |
| *Hydrocodone* | *CYP2D6* | | | | *Pain Management* |
| *Methadone** | *CYP2D6* | *CYP2B6* | | | *Pain Management* |
| *Oxycodone** | *CYP2D6* | | | | *Pain Management* |
| *Tramadol* | *CYP2D6* | | | | *Pain Management* |
| *Capecitabine* | *DPYD* | | | | *Hematology / Oncology* |
| *Fluorouracil* | *DPYD* | | | | *Hematology / Oncology* |
| *Tegafur** | *DPYD* | | | | *Hematology / Oncology* |
| *Ondansetron* | *CYP2D6* | | | | *Gastroenterology* |
| *Tropisetron* | *CYP2D6* | | | | *Gastroenterology* |
| *Fosphenytoin* | *HLA-B*15:02* | *CYP2C9* | | | *Neurology* |
| *Phenytoin* | *HLA-B*15:02* | *CYP2C9* | | | *Neurology* |

| | | | | | |
|---|---|---|---|---|---|
| *Tacrolimus* | *CYP3A5* | | | | *Transplant* |
| *Azathioprine* | *TPMT* | *NUDT15* | | | *Rheumatology / Transplant* |
| *Mercaptopurine* | *TPMT* | *NUDT15* | | | *Hematology / Oncology* |
| *Thioguanine* | *TPMT* | *NUDT15* | | | *Hematology / Oncology* |
| *Amitriptyline* | *CYP2D6* | *CYP2C19* | | | *Behavioral Health* |
| *Clomipramine* | *CYP2D6* | *CYP2C19* | | | *Behavioral Health* |
| *Desipramine* | *CYP2D6* | | | | *Behavioral Health* |
| *Doxepin* | *CYP2D6* | *CYP2C19* | | | *Behavioral Health* |
| *Imipramine* | *CYP2D6* | *CYP2C19* | | | *Behavioral Health* |
| *Nortriptyline* | *CYP2D6* | | | | *Behavioral Health* |
| *Trimipramine* | *CYP2D6* | *CYP2C19* | | | *Behavioral Health* |
| *Voriconazole* | *CYP2C19* | | | | *Infectious Diseases* |
| *Warfarin*[a] | *CYP2C9* | *VKORC1* | *CYP4F2* | *CYP2C rs12777823* | *Cardiology* |

** CPIC level C (no recommendation)*
*[a] Warfarin and the genes only relevant for warfarin are displayed with a static "yellow" warning; the annotation states that further information is needed to calculate the right dose*
