## Supplementary Material 6 for "Design and Implementation of PharMe: A Mobile Application to Return Pharmacogenomic Test Results to Patients"

### Amitriptyline

Pharmacogenomic (PGx) Report

*Disclaimer: The information contained in this PDF document is intended solely for use by trained health care professionals. It is provided for informational purposes only and should not be considered medical advice. The content may include technical terminology and clinical data that are intended for professional interpretation and application. Recipients are advised to exercise professional judgment and discretion when utilizing the information contained herein. If you are not a trained health care professional, please consult with a qualified medical practitioner or specialist before interpreting or applying the information provided in this document.*

**Medication type:** Tricyclic antidepressant

**Indication:** Amitriptyline is used to treat depression. It also can be used to treat pain.

**Brand names:** Elavil

#### User data

**Genotype:** CYP2D6: *1/*4, CYP2C19: *1/*1

**Phenotype:** CYP2D6: Intermediate Metabolizer, CYP2C19: Normal Metabolizer

**Activity score:** CYP2D6: 1.0, CYP2C19: Normal Metabolizer

**Tested alleles:** CYP2D6: *1(reference), *2, *3, *4, *4.018, *4M, *4N, *5 (gene deletion), *6, *7, *8, *9, *10, *11, *12, *13, *14, *15, *17, *18, *19.001,*19.002, *29, *31, *34,*35, *36, *39, *40, *41, *42, *49, *53, *56A, *56B, *59, *64, *68, *69, *109, *114; *119, duplication (x2), multiplication (x3), CYP2C19: *1(reference), *2, *3, *4A, *4B, *5, *6, *7, *8, *9, *10, *17, *35

**User guideline:** ⚠️ You break down amitriptyline slower than expected. You have an increased risk for side effects. Based on your genes, amitriptyline may be used at a lower dose. Consult your pharmacist or doctor for more information.

#### Clinical guideline(s)

*For more fine-grained information please refer to the original guideline(s) by following the URL(s) below.*

---

**CYP2D6, CYP2C19 and Tricyclic Antidepressants**

**CPIC guideline link:**
[https://cpicpgx.org/guidelines/guideline-for-tricyclic-antidepressants-and-cyp2d6-and-cyp2c19/](https://cpicpgx.org/guidelines/guideline-for-tricyclic-antidepressants-and-cyp2d6-and-cyp2c19/)

**CPIC recommendation:** Consider a 25% reduction of recommended starting dose. Utilize therapeutic drug monitoring to guide dose adjustments.

**CPIC implication for CYP2D6:** Reduced metabolism of TCAs to less active compounds compared to normal metabolizers; Higher plasma concentrations of active drug will increase the probability of side effects
**CPIC implication for CYP2C19:** Normal metabolism of tertiary amines

**CPIC comment:** Patients may receive an initial low dose of a tricyclic, which is then increased over several days to the recommended steady-state dose. The starting dose in this guideline refers to the recommended steady-state dose. Dosing recommendations only apply to higher initial doses of TCAs for treatment of conditions such as depression. See other considerations for dosing recommendations for conditions where lower initial doses are used, such as neuropathic pain.

### Amitriptyline

Pharmacogenomic (PGx) Report

*Disclaimer: The information contained in this PDF document is intended solely for use by trained health care professionals. It is provided for informational purposes only and should not be considered medical advice. The content may include technical terminology and clinical data that are intended for professional interpretation and application. Recipients are advised to exercise professional judgment and discretion when utilizing the information contained herein. If you are not a trained health care professional, please consult with a qualified medical practitioner or specialist before interpreting or applying the information provided in this document.*

**Medication type:** Tricyclic antidepressant

**Indication:** Amitriptyline is used to treat depression. It also can be used to treat pain.

**Brand names:** Elavil

#### User data

**Genotype:** CYP2D6: *1/*4, CYP2C19: *1/*1

**Phenotype:** CYP2D6: Poor Metabolizer*, CYP2C19: Normal Metabolizer

* The user's CYP2D6 result was changed from "Intermediate Metabolizer" to "Poor Metabolizer"; this is because the user is currently taking one or more medications that strongly slow down (inhibit) the activity of CYP2D6 (see below).

Strong CYP2D6 inhibitor: Fluoxetine (brand names: Act Fluoxetine, Prozac, Sarafem, Symbyax)

**Activity score:** CYP2D6: 0.0 (1.0 adjusted based on current medications, see phenotype), CYP2C19: Normal Metabolizer

**Tested alleles:** CYP2D6: *1(reference), *2, *3, *4, *4.018, *4M, *4N, *5 (gene deletion), *6, *7, *8, *9, *10, *11, *12, *13, *14, *15, *17, *18, *19.001,*19.002, *29, *31, *34,*35, *36, *39, *40, *41, *42, *49, *53, *56A, *56B, *59, *64, *68, *69, *109, *114; *119, duplication (x2), multiplication (x3), CYP2C19: *1(reference), *2, *3, *4A, *4B, *5, *6, *7, *8, *9, *10, *17, *35

**User guideline:** ❌ You break down amitriptyline much slower than expected. You have an increased risk for side effects. Based on your genes, amitriptyline may not be the right medication for you. However, if needed, amitriptyline may be used at a lower dose. Consult your pharmacist or doctor for more information.

#### Clinical guideline(s)

*For more fine-grained information please refer to the original guideline(s) by following the URL(s) below.*

**CYP2D6, CYP2C19 and Tricyclic Antidepressants**

**CPIC guideline link:** https://cpicpgx.org/guidelines/guideline-for-tricyclic-antidepressants-and-cyp2d6-and-cyp2c19/

**CPIC recommendation:** Avoid amitriptyline use. If a amitripyline is warranted, consider a 50% reduction of recommended starting dose. Utilize therapeutic drug monitoring to guide dose adjustments.

**CPIC implication for CYP2D6:** Greatly reduced metabolism of TCAs to less active compounds compared to normal metabolizers; Higher plasma concentrations of active drug will increase the probability of side effects
**CPIC implication for CYP2C19:** Normal metabolism of tertiary amines

**CPIC comment:** Patients may receive an initial low dose of a tricyclic, which is then increased over several days to the recommended steady-state dose. The starting dose in this guideline refers to the recommended steady-state dose. Dosing recommendations only apply to higher initial doses of TCAs for treatment of conditions such as depression. See other considerations for dosing recommendations for conditions where lower initial doses are used, such as neuropathic pain.
